# Evaluation of Women with Peripartum or Dilated Cardiomyopathy and Their First-Degree Relatives: The DCM Precision Medicine Study

**DOI:** 10.1101/2025.02.18.25322501

**Authors:** Evan P. Kransdorf, Rashmi Jain, Jonathan O. Mead, Garrie Haas, Mark Hofmeyer, Gregory A. Ewald, Jamie Diamond, Anjali Owens, Brian Lowes, Douglas Stoller, W. H. Wilson Tang, Mark H. Drazner, Cindy M. Martin, Palak Shah, Jose Tallaj, Stuart Katz, Javier Jimenez, Supriya Shore, Frank Smart, Jessica Wang, Stephen S. Gottlieb, Daniel P. Judge, Gordon S. Huggins, Jason Cowan, Patricia Parker, Jinwen Cao, Natalie S. Hurst, Elizabeth Jordan, Hanyu Ni, Daniel D. Kinnamon, Ray E. Hershberger

## Abstract

**Background:** Rare variant genetics have been associated with peripartum cardiomyopathy (PPCM) but the role of genetics remains unsettled.

**Objective:** The study sought to compare dilated cardiomyopathy (DCM) genetic risk in first-degree relatives (FDRs) of female patients with DCM or PPCM (probands), and to assess DCM-relevant rare variant prevalence in DCM/PPCM probands and population controls.

**Methods:** Clinical and genetic data were analyzed from the DCM Precision Medicine Study. Risk of DCM or partial DCM, where pDCM was defined as left ventricular (LV) enlargement or a LV ejection fraction of <50%, was estimated in 665 FDRs from 452 female probands, all of whom had been pregnant, of which 67 had PPCM and 385 had DCM; prevalence of pathogenic, likely pathogenic or uncertain significance variants (P/LP/VUS) was estimated among probands.

**Results:** The risk of DCM/pDCM for FDRs of PPCM probands was similar to that for FDRs of DCM probands (HR, 0.77; 95% CI, 0.47 – 1.28). Estimated DCM prevalence among the lowest-risk FDRs of non-Hispanic EA probands with PPCM (7.0% [95% CI, 0%-14.1%] females, 9.0% [95% CI, 1.6%-16.3%] males) exceeded population estimates from a UK Biobank study (0.30% females, 0.63% males). Estimated prevalences of a P/LP/VUS among AA and EA probands with PPCM were 55.4% (95% CI, 33.1%-77.7%) and 66.0% (95% CI, 38.6%-93.3%), respectively. The estimated prevalence of P/LP variants among EA PPCM probands (26.6%; 95% CI, 12.6%-40.6%) exceeded a population estimate from a UK Biobank study (0.6%).

**Conclusion:** The risk of DCM/pDCM among FDRs was similar regardless of whether their probands had PPCM or DCM. Also, DCM-relevant rare variant findings for females with PPCM or DCM were similar and greater than in population controls suggesting a shared genetic basis for PPCM and DCM. These findings underscore the need for genetic evaluations in all PPCM patients.

**Condensed Abstract:** This is the first study to assess and compare dilated cardiomyopathy (DCM) risk in first-degree relatives (FDRs) of females with peripartum cardiomyopathy (PPCM) and (DCM). The similar risk of DCM or a partial phenotype of DCM in FDRs of females with PPCM and DCM suggests a common genetic contribution to PPCM and DCM. Also, the prevalence of DCM-relevant rare genetic variants was similar between FDRs of females probands diagnosed with PPCM and DCM within European and African ancestry groups and much higher than in population controls. These findings underscore the need of a genetics evaluation for all females with PPCM.

**Clinical Trial:** clinicaltrials.gov, NCT03037632

## Introduction

Peripartum cardiomyopathy (PPCM) is defined as left ventricular (LV) systolic dysfunction that presents with heart failure during pregnancy or in the postpartum period without other clinically identifiable cardiovascular causes such as coronary or valvular disease.^1,2^ PPCM is a significant contributor to maternal mortality.^3^ Previously identified risk factors for PPCM include Black race, pre-eclampsia, hypertension, advanced maternal age, and multi-gestational pregnancies.^1,2^ The underlying cause of PPCM remains incompletely defined with alternative hypotheses suggesting genetic and/or pregnancy-related clinical causes.^2^ A genetic contribution to PPCM has been suggested based on familial clustering of PPCM.^4–6^ Also, rare variants in genes associated with dilated cardiomyopathy (DCM) have been identified in females with PPCM,^5,7–9^ including evidence that equivalent fractions of females with PPCM and DCM have DCM-relevant genetic findings.^7,8^ However, whether genetics is only a contributing factor or constitutes a primary causal basis for PPCM has not been resolved.

To address this question we analyzed data from the family-based DCM Precision Medicine Study with a central hypothesis that most of DCM has a genetic basis.^10^ The study enrolled 1220 DCM probands (patients), of which 44% were female, and 1693 first-degree relatives (FDRs).^11^ The FDRs provided a unique opportunity to evaluate the role of genetics in females with PPCM as compared with those with DCM, as this study postulated that if PPCM shared a common genetic basis with DCM, the risk of DCM or a partial DCM (pDCM) phenotype among the FDRs of females with PPCM would be similar to that in FDRs of females with DCM. Conversely, if DCM-relevant genetic risk factors did not contribute to PPCM, the FDRs of females with PPCM would have a much lower risk of DCM and pDCM. This was tested by evaluating the FDRs of females probands from the DCM Precision Medicine Study who had been pregnant at least once with PPCM or DCM. The genetic findings for females with PPCM were also compared to DCM and to general population estimates from the UK biobank study.

## Methods

### Participants

The DCM Precision Medicine study was a cross-sectional study of families at 25 U.S. clinical sites; patients with DCM (probands) and FDRs were enrolled between June 2016 and April 2021.^10,11^ All probands met criteria for DCM, defined as LV systolic dysfunction (LVSD; LV ejection fraction <50%) and LV enlargement (LVE) with other identifiable clinical causes, except genetic, excluded.^10,11^ Cardiac magnetic resonance imaging data when available validated DCM diagnoses.^12^ The Institutional Review Boards (IRB) at the Ohio State University and all clinical sites approved the initial study, followed by single IRB oversight at the University of Pennsylvania. All participants gave written informed consent.

Probands assigned as female at birth and with a history of pregnancy were included in this analysis (Figure 1). PPCM cases were females that met the above definition of DCM during pregnancy or within five months of the post-partum period. All other female probands were assigned to the DCM group for comparison.

**Figure 1.**
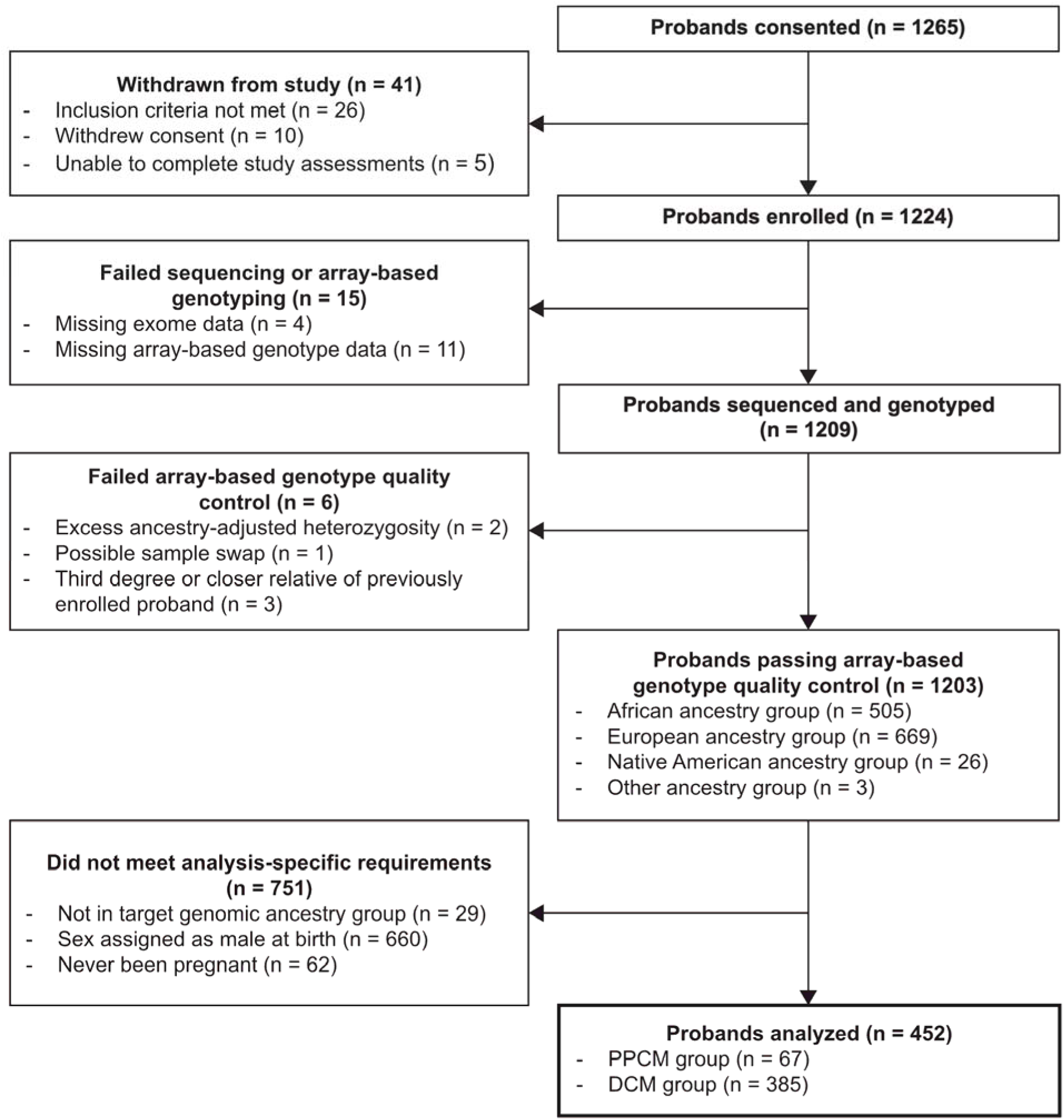
Selection of DCM Precision Medicine Study participants for analysis

### Data Collection

Clinical data were collected and centrally adjudicated to confirm DCM in probands, and DCM, pDCM, or neither in FDRs, as previously described.^11,13^ pDCM was defined in FDRs as LV enlargement or a LV ejection fraction <50%, as previously defined.^11^ Participant demographic information, health history, and cardiovascular clinical data were obtained through structured interviews and review of medical records, including data for all females regarding pregnancy history.^10,11,13^ Research exome sequencing of probands was conducted and data processed as described previously.^14,15^ Rare protein-altering variants in 36 genes considered relevant for DCM at the development of the study,^10^ including the 19 genes classified as definitive, strong and moderate evidence by the Clinical Genome Resource (ClinGen),^16^ were identified and interpreted according to American College of Medical Genetics/Association for Molecular Pathology (ACMG/AMP) and Clinical Genome Resource (ClinGen)-based criteria tailored to DCM.^14,15^

The DCM Consortium is aware of issues for the collection, analysis, presentation, and discussion of race, ethnicity, and ancestry and has adopted recommended approaches.^17^ Self-reported race and ethnicity data were included in this study because of their relevance for health outcomes; they were self-reported by participants using structured race (Native American or Alaska Native, Asian, African American, Native Hawaiian or Pacific Islander, White, more than 1 race, or unknown) and Hispanic ethnicity (yes, no, or unknown) categories. Genomic ancestry proportions in probands were estimated from Illumina Global Screening Array genotypes and used to assign individuals to groups based on predominant ancestry as previously described.^15^ Individuals not in the African ancestry (AA) or European ancestry (EA) group were not analyzed due to small numbers (n = 29).

### Statistical Analysis

Detailed statistical methods are provided in eAppendix 1. All statistical analyses were performed in SAS/STAT version 15.2 software, version 9.4 (TS1M7) of the SAS System for 64-bit Linux (SAS Institute Inc) and R version 4.2.2 (R Foundation, Vienna, Austria). Statistical tests and confidence intervals were two-sided with α = 0.05 unless otherwise noted.

Differences in the distributions of demographic and clinical characteristics between females in the PPCM and DCM groups within proband enrollment sites were assessed using a generalized Cochran-Mantel-Haenszel test^18,19^ (categorical) and the Boos-Brownie test^20^ of the relative effect (continuous), both of which treat sites as random.

To estimate ancestry-specific prevalences of PPCM for a proband at a typical US advanced heart failure program, a generalized linear mixed model for a logit-linked binary outcome (PPCM or DCM) with covariates for ancestry group (AA or EA), ethnicity (Hispanic or non-Hispanic), and enrollment age quartile was fit to the proband data. The model included a random intercept to adjust for enrollment site heterogeneity. Marginally standardized prevalence estimates at a typical US advanced heart failure program were calculated for probands of AA and EA as described in eAppendix 1.

To investigate the potential association between PPCM status and DCM-relevant rare variant findings, trinomial outcomes were defined for each proband based on the number of variants classified as Pathogenic, Likely Pathogenic or Variant of Uncertain Significance (P/LP/VUS; 0, 1, or >1) and the most deleterious variant identified (none, VUS, or P/LP). These outcomes were modeled using a previously published hierarchical logit mixed model^15^ with PPCM status, ancestry group, ethnicity, and quartile of age at DCM diagnosis as fixed effects and proband enrollment site random effects to account for potential site heterogeneity. The Morel-Bokossa-Neerchal bias-corrected empirical covariance estimator with sites as independent units was used to counter the well-known downward bias in model-based covariance estimates.^21,22^ In addition to odds ratios comparing groups, this model fit was used to obtain marginally standardized estimates of outcome probabilities at a typical US advanced heart failure program, as described in eAppendix 1. In addition, a logistic regression model with PPCM status, ancestry group, ethnicity), and quartile of age at DCM diagnosis fixed effects was used to estimate the prevalence of P/LP findings in the population of PPCM probands for comparison with a general population prevalence estimate from a prior UK Biobank study,^23^ as described in eAppendix 1.

Variants were grouped by the PPCM classification of the study proband(s) in whom they were identified (PPCM, DCM, or both). Variant characteristics were compared using the exact Pearson Χ^2^ test for nominal variables and the Kruskal-Wallis test for continuous variables. For post hoc pairwise comparisons between PPCM classifications, Holm-Bonferroni corrected exact Monte-Carlo p-values for the Χ^2^ test involving only those 2 groups or Dwass-Steel-Critchlow-Fligner multiplicity-adjusted p-values based on pairwise Wilcoxon rank sum tests were used.^24,25^ The exact Monte Carlo p-value estimate based on 100,000 replicates was used for the Pearson Χ^2^ test.

Data on the DCM or partial DCM status of each enrolled FDR at enrollment was used to model age-specific cumulative risks using a previously published approach.^11^ In this approach, the unobserved age at disease onset in FDRs was assumed to have a marginal distribution with a Weibull baseline survivor function influenced by covariates and site random effects through a proportional hazards model. Covariates included proband PPCM status, ancestry group, ethnicity, and age at diagnosis quartile and first-degree relative biological sex. This model was fit as a generalized estimating equation-type generalized linear mixed model^22,26^ with a binary outcome (presence or absence of DCM or DCM/pDCM), complementary log-log link, and working independence correlation structure using residual subject specific pseudolikelihood; intra-familial correlation was accounted for by using the Morel-Bokossa-Neerchal bias-corrected empirical covariance estimator with sites as independent units.^21^ In addition to conditional hazard ratios comparing groups at a particular site, this model fit was used to obtain estimates of age-specific cumulative risk among the lowest-risk FDRs of non-Hispanic EA probands with PPCM or DCM at a typical US advanced heart failure program and to estimate DCM prevalence among such FDRs of PPCM patients seen at these programs for comparison to general population DCM prevalence estimates from a prior UK Biobank study,^27^ as detailed in eAppendix 1.

## Results

Of 1203 probands, 452 female probands with ≥1 pregnancy were evaluated (Figure 1), including 67 with PPCM and 385 with DCM. The principal differences observed between females with PPCM and DCM were an earlier age of diagnosis and earlier age of study enrollment (Table 1). There was a higher percentage of Black (self-identified) probands in the PPCM group, but these differences were not statistically significant.

**Table 1.** Demographic and clinical characteristics of female probands with DCM, by PPCM classification.

|  | **PPCM**<br>**N = 67** | **DCM**<br>**N = 385** | **P value<sup>b</sup>** |
| --- | --- | --- | --- |
| **Characteristic** |  |  |  |
| Predominant genomic ancestry,<sup>a</sup> No. (%) |  |  |  |
| African | 38 (56.7) | 167 (43.4) | 0.33 |
| European | 29 (43.3) | 218 (56.6) |  |
| Self-identified race, No. (%) |  |  |  |
| Black | 38 (56.7) | 171 (44.4) | 0.43 |
| White | 29 (43.3) | 213 (55.3) |  |
| More than one race | 0 (0.0) | 1 (0.3) |  |
| Continental ancestry proportion, mean (SD) |  |  |  |
| African | 0.48 (0.41) | 0.37 (0.40) | 0.15 |
| European | 0.47 (0.40) | 0.59 (0.39) | 0.28 |
| Native American | 0.02 (0.07) | 0.02 (0.06) | 0.43 |
| East Asian | 0.01 (0.01) | 0.01 (0.03) | 0.18 |
| South Asian | 0.02 (0.03) | 0.02 (0.02) | 0.001 |
| Self-identified Hispanic ethnicity, No. (%) | 6 (9.0) | 20 (5.2) | 0.19 |
| Age at enrollment, years, median (IQR) | 41.6 (33.1 – 48.6) | 56.2 (46.2 – 64.5) | <0.001 |
| Age at diagnosis, years, median (IQR) | 29.7 (24.4 – 34.5) | 47.4 (39.8 – 55.6) | <0.001 |
| BMI, median (IQR) | 29.8 (26.2 – 36.3) | 30.3 (26.1 – 35.7) | 0.40 |
| Comorbidities, No. (%) |  |  |  |
| Hypertension | 28 (41.8) | 202 (52.5) | 0.051 |
| High cholesterol | 8 (11.9) | 110 (28.6) | 0.005 |
| Diabetes | 15 (22.4) | 92 (23.9) | 0.57 |
| Cancer | 1 (1.5) | 17 (4.4) | 0.33 |
| Lung disease | 2 (3.0) | 25 (6.5) | 0.32 |
| Asthma | 11 (16.4) | 75 (19.5) | 0.32 |
| Presenting symptoms, No. / No. avail. (%) |  |  |  |
| Dyspnea on exertion | 26 / 66 (39.4) | 166 / 384 (43.2) | 0.76 |
| Edema | 19 / 66 (28.8) | 104 / 384 (27.1) | 0.45 |
| Orthopnea | 17 / 66 (25.8) | 105 / 384 (27.3) | 0.69 |
| Weight gain | 13 / 66 (19.7) | 76 / 384 (19.8) | 0.87 |
| Fatigue | 25 / 66 (37.9) | 164 / 384 (42.7) | 0.26 |
| Palpitations | 20 / 66 (30.3) | 99 / 384 (25.8) | 0.35 |
| Syncope | 4 / 66 (6.1) | 37 / 384 (9.6) | 0.25 |
| Arrhythmia | 10 / 66 (15.2) | 62 / 384 (16.2) | 0.85 |
| Asymptomatic | 0 / 66 (0.0) | 19 / 384 (5.0) | 0.02 |
| Blood pressure measures, median (IQR)<br>[No. avail.]<sup>c</sup> |  |  |  |
| Systolic (mmHg) | 112 (100 – 120) [55] | 114 (103 – 126) [319] | 0.51 |
| Diastolic (mmHg) | 72 (63 – 78) [55] | 70 (63 – 77) [319] | 0.80 |
| Mean arterial (mmHg) | 85 (77 – 92) [55] | 85 (77 – 93) [319] | 0.88 |
| Left ventricular ejection fraction (%),<br>median (IQR) [No. avail.]<sup>d</sup> | 23.0 (17.5 – 32.5) [66] | 22.5 (16.0 – 30.0) | 0.79 |
| Left ventricular internal diastolic<br>dimension (mm), median (IQR)<br>[No. avail.]<sup>d</sup> | 61.0 (57.1 – 70.0) [66] | 61.0 (57.0 – 68.0) | 0.52 |
| Left ventricular internal diastolic<br>dimension (z-score), median (IQR)<br>[No. avail.]<sup>d,e</sup> | 4.4 (3.3 – 6.1) [66] | 4.4 (3.2 – 5.9) | 0.64 |
| Interventions, No. / No avail. (%)<sup>d</sup> |  |  |  |
| Left ventricular assist device | 10 (14.9) | 66 (17.1) | 0.91 |
| Heart transplant | 8 (11.9) | 40 (10.4) | 0.51 |
| Bi-ventricular pacemaker | 5 / 66 (7.6) | 67 / 371 (18.1) | 0.13 |
| Implantable cardioverter<br>defibrillator | 42 / 65 (64.6) | 255 / 383 (66.6) | 0.58 |

BMI = body mass index; DCM = dilated cardiomyopathy; IQR = interquartile range; PPCM = peripartum cardiomyopathy; SD = standard deviation.
<sup>a</sup> Genomic ancestry was determined based on global ancestry proportions inferred from array-based genotypes; an individual's predominant ancestry group was defined as the continental ancestry group (African, East Asian, European, Native American, or South Asian) accounting for the highest proportion of their genomic ancestry. Individuals with predominant continental ancestry other than African or European ancestry were not analyzed due to small numbers (see Methods).
<sup>b</sup> For categorical variables, two-sided p-values are from a generalized Cochran-Mantel-Haenszel test using the general association statistic, which is designed to detect within-site differences in the distribution of the row variable between PPCM and DCM that are consistent across sites. For continuous variables, two-sided p-values are from the Boos-Brownie test of the relative effect, which is designed to detect whether values in the PPCM group are systematically higher or lower than in the DCM group consistently across sites. Both tests use proband enrollment sites as the primary sampling units.
<sup>c</sup> Does not include measurements recorded after left ventricular assist device placement.
<sup>d</sup> The number available is not provided if it equates to the sample size defined in the column header.
<sup>e</sup> Calculated based on sex and height for all probands with heights of at least 137 cm.

From the 452 female probands, 665 FDRs were available for analysis (Table 2). The demographics of both groups were similar regardless of their proband’s assignment as PPCM or DCM.

**Table 2.**
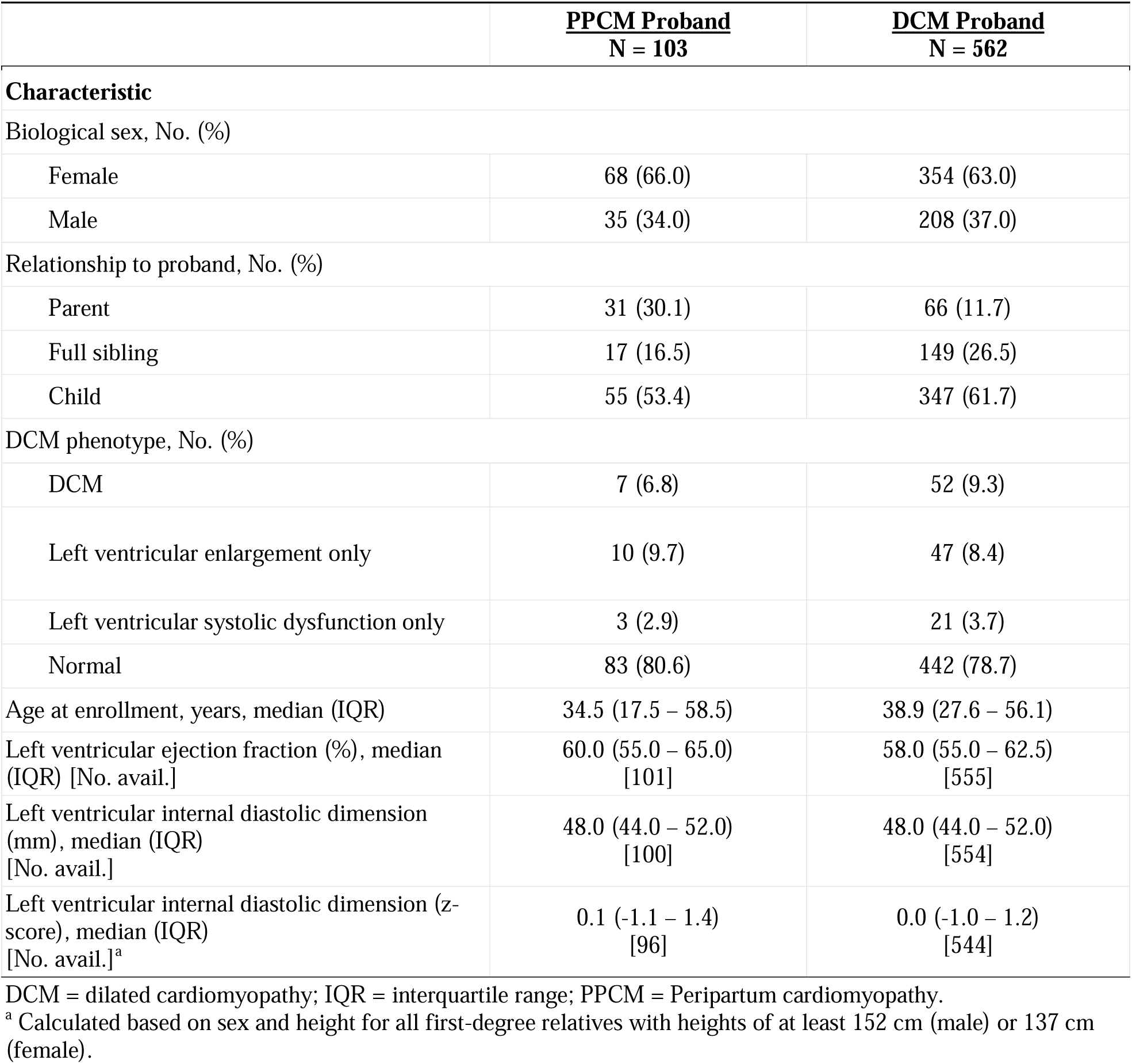
Demographic and clinical characteristics of first-degree relatives of female probands with DCM, by proband PPCM classification.

|  | **<u>PPCM Proband</u>**<br>**N = 103** | **<u>DCM Proband</u>**<br>**N = 562** |
| --- | --- | --- |
| **Characteristic** |  |  |
| Biological sex, No. (%) |  |  |
| Female | 68 (66.0) | 354 (63.0) |
| Male | 35 (34.0) | 208 (37.0) |
| Relationship to proband, No. (%) |  |  |
| Parent | 31 (30.1) | 66 (11.7) |
| Full sibling | 17 (16.5) | 149 (26.5) |
| Child | 55 (53.4) | 347 (61.7) |
| DCM phenotype, No. (%) |  |  |
| DCM | 7 (6.8) | 52 (9.3) |
| Left ventricular enlargement only | 10 (9.7) | 47 (8.4) |
| Left ventricular systolic dysfunction only | 3 (2.9) | 21 (3.7) |
| Normal | 83 (80.6) | 442 (78.7) |
| Age at enrollment, years, median (IQR) | 34.5 (17.5 – 58.5) | 38.9 (27.6 – 56.1) |
| Left ventricular ejection fraction (%), median (IQR) [No. avail.] | 60.0 (55.0 – 65.0)<br>[101] | 58.0 (55.0 – 62.5)<br>[555] |
| Left ventricular internal diastolic dimension (mm), median (IQR) [No. avail.] | 48.0 (44.0 – 52.0)<br>[100] | 48.0 (44.0 – 52.0)<br>[554] |
| Left ventricular internal diastolic dimension (z-score), median (IQR) [No. avail.]<sup>a</sup> | 0.1 (-1.1 – 1.4)<br>[96] | 0.0 (-1.0 – 1.2)<br>[544] |

DCM = dilated cardiomyopathy; IQR = interquartile range; PPCM = Peripartum cardiomyopathy.
<sup>a</sup> Calculated based on sex and height for all first-degree relatives with heights of at least 152 cm (male) or 137 cm (female).

### Ancestry-specific PPCM prevalence estimates in probands

The prevalence of PPCM was estimated separately for AA and EA probands using the model in eTable 1. Among AA probands the estimated prevalence of PPCM was 16.8% (95% CI, 11.1% - 22.6%). For a similar population of EA probands, the estimated prevalence was 14.4% (95% CI, 7.6% - 21.2%). The mean difference in PPCM prevalence between ancestries was 2.4% (95% CI, −6.8% - 11.6%), with no statistically significant difference.

### Genetic findings in probands with PPCM versus DCM

A detailed tabulation of P/LP/VUS variants by gene and PPCM status of the proband(s) in whom they were found is provided (eTable 2). The model-based estimates of the prevalence of harboring a variant classified as P/LP/VUS among AA and EA probands with PPCM at a typical US heart failure program were 55.4% (95% CI, 33.1%-77.7%) and 66.0% (95% CI, 38.6%-93.3%), respectively (Table 3). These prevalences were similar among probands with DCM within each ancestry (Table 3), reflecting similar odds of P/LP/VUS findings in probands with PPCM or DCM (OR, 0.79; 95% CI, 0.25-2.49; Table 4).

**Table 3.** Variant classification results in female probands, by ancestry and PPCM classification.

| <u>Outcome</u> | <u>African</u><br>N = 205 |  |  |  | <u>European</u><br>N = 247 |  |  |  |
| --- | --- | --- | --- | --- | --- | --- | --- | --- |
|  | <u>PPCM</u><br>N = 38 |  | <u>DCM</u><br>N = 167 |  | <u>PPCM</u><br>N = 29 |  | <u>DCM</u><br>N = 218 |  |
|  | <u>Crude</u> | <u>Model-based<sup>a</sup></u> | <u>Crude</u> | <u>Model-based<sup>a</sup></u> | <u>Crude</u> | <u>Model-based<sup>a</sup></u> | <u>Crude</u> | <u>Model-based<sup>a</sup></u> |
|  | No. (%) | % (Joint 95% CI) | No. (%) | % (Joint 95% CI) | No. (%) | % (Joint 95% CI) | No. (%) | % (Joint 95% CI) |
| <u>Most deleterious variant identified</u> |  |  |  |  |  |  |  |  |
| None | 15 (39.5) | 44.6 (22.3 – 66.9) | 77 (46.1) | 38.9 (19.7 – 58.2) | 11 (37.9) | 34.0 (6.7 – 61.4) | 77 (35.3) | 29.0 (18.5 – 39.6) |
| VUS | 19 (50.0) | 47.7 (25.2 – 70.1) | 78 (46.7) | 53.3 (32.5 – 74.1) | 10 (34.5) | 36.4 (7.9 – 64.9) | 80 (36.7) | 41.2 (27.5 – 54.8) |
| P/LP | 4 (10.5) | 7.8 (0.0 – 15.7) | 12 (7.2) | 7.8 (1.1 – 14.4) | 8 (27.6) | 29.5 (12.5 – 46.5) | 61 (28.0) | 29.8 (15.5 – 44.2) |
| <u>Number of P/LP/VUS variants identified</u> |  |  |  |  |  |  |  |  |
| None | 15 (39.5) | 44.6 (22.4 – 66.7) | 77 (46.1) | 38.9 (19.8 – 58.1) | 11 (37.9) | 34.0 (6.8 – 61.3) | 77 (35.3) | 29.0 (18.6 – 39.4) |
| 1 | 18 (47.4) | 43.2 (19.2 – 67.3) | 63 (37.7) | 41.4 (24.8 – 58.0) | 13 (44.8) | 47.8 (16.0 – 79.7) | 90 (41.3) | 43.3 (31.6 – 55.0) |
| >1 | 5 (13.2) | 12.2 (1.4 – 23.0) | 27 (16.2) | 19.7 (9.2 – 30.1) | 5 (17.2) | 18.1 (4.3 – 32.0) | 51 (23.4) | 27.7 (17.2 – 38.2) |

CI = confidence interval; DCM = dilated cardiomyopathy; LP = likely pathogenic; P = pathogenic; PPCM = peripartum cardiomyopathy; VUS = variant of uncertain significance.
<sup>a</sup>Estimates from hierarchical logit mixed models for trinomial outcomes presented in Table 4 and the Morel-Bokossa-Neerchal bias-corrected estimate of their empirical covariance matrix with sites as independent units were used to obtain marginally standardized estimates of and delta method joint 95% CIs for outcome probabilities for female probands in a particular ancestry group and PPCM classification. All CIs were produced using the standard normal distribution and the Bonferroni correction over the 3 possible outcome categories. Each patient subpopulation defined by ancestry group and PPCM status was assumed to be balanced across the two youngest age quartiles ( $\leq 44.47$ years of age) with a proportion of Hispanics equal to the 2021 US census population estimate (18.9%).<sup>40</sup> As evidence of site heterogeneity was found in both models, these estimates can be interpreted as applicable to patients seen at a typical US advanced heart failure program, defined as a program at the mean or mode of the random effects distribution describing the population of such programs.

**Table 4.** Associations of PPCM classification, ancestry, ethnicity, sex, and age at diagnosis with variant classification results in probands.

| Predictor | Most Deleterious Variant |  |  |  | Number of Variants |  |  |  |
| --- | --- | --- | --- | --- | --- | --- | --- | --- |
|  | P/LP/VUS vs. None |  | P/LP vs. VUS<br>Given P/LP/VUS |  | >=1 P/LP/VUS vs. None |  | >1 Variant vs. 1 Variant<br>Given >=1 P/LP/VUS |  |
|  | Odds Ratio<br>(95% CI)<sup>a,b</sup> | P<sup>a</sup> | Odds Ratio<br>(95% CI)<sup>a,c</sup> | P<sup>a</sup> | Odds Ratio<br>(95% CI)<sup>a,b</sup> | P<sup>a</sup> | Odds Ratio<br>(95% CI)<sup>a,c</sup> | P<sup>a</sup> |
| <u>PPCM classification</u> |  |  |  |  |  |  |  |  |
| PPCM | 0.79 (0.25 – 2.49) | 0.69 | 1.12 (0.39 – 3.24) | 0.84 | 0.79 (0.25 – 2.48) | 0.69 | 0.59 (0.19 – 1.85) | 0.37 |
| DCM | 1.00 | - | 1.00 | - | 1.00 | - | 1.00 | - |
| <u>Ancestry group</u> |  |  |  |  |  |  |  |  |
| African | 0.64 (0.37 – 1.12) | 0.12 | 0.20 (0.09 – 0.44) | <0.001 | 0.64 (0.37 – 1.12) | 0.12 | 0.74 (0.44 – 1.25) | 0.26 |
| European | 1.00 | - | 1.00 | - | 1.00 | - | 1.00 | - |
| <u>Self-reported ethnicity</u> |  |  |  |  |  |  |  |  |
| Hispanic | 1.11 (0.35 – 3.57) | 0.86 | 0.76 (0.23 – 2.50) | 0.66 | 1.11 (0.35 – 3.54) | 0.86 | 1.47 (0.48 – 4.53) | 0.50 |
| Non-Hispanic | 1.00 | - | 1.00 | - | 1.00 | - | 1.00 | - |
| <u>Diagnosis age quartile</u> |  |  |  |  |  |  |  |  |
| I: [9.78, 34.99] | 1.00 | - | 1.00 | - | 1.00 | - | 1.00 | - |
| II: [35.01, 44.47] | 0.82 (0.43 – 1.58) | 0.55 | 0.75 (0.32 – 1.73) | 0.50 | 0.82 (0.43 – 1.56) | 0.55 | 0.89 (0.34 – 2.36) | 0.82 |
| III: [44.48, 54.09] | 0.65 (0.32 – 1.34) | 0.25 | 0.89 (0.32 – 2.45) | 0.83 | 0.65 (0.32 – 1.33) | 0.24 | 0.72 (0.28 – 1.82) | 0.49 |
| IV: [54.21, 78.31] | 0.50 (0.27 – 0.95) | 0.03 | 1.00 (0.33 – 3.05) | 1.00 | 0.50 (0.27 – 0.94) | 0.03 | 1.05 (0.35 – 3.12) | 0.93 |

CI = confidence interval; DCM = dilated cardiomyopathy; LP = likely pathogenic; P = pathogenic; PPCM = peripartum cardiomyopathy; VUS = variant of uncertain significance.
<sup>a</sup> Odds ratios and Wald 95% CIs from hierarchical logit mixed models for trinomial outcomes based on the most deleterious P/LP/VUS variant identified (none, VUS, or P/LP) and the number of P/LP/VUS variants identified (0, 1, or >1) were adjusted for all other variables shown in the table. Two-sided Wald p-values are for the null hypothesis that the odds ratio is 1. All p-values and CIs were produced using the Morel-Bokossa-Neerchal bias-corrected estimate of the empirical covariance matrix and the standard normal distribution.
<sup>b</sup> As evidence of site heterogeneity was found, these estimates can be interpreted as comparing DCM patients within the same US advanced heart failure program.
<sup>c</sup> As no evidence of site heterogeneity was found, these estimates can be interpreted as comparing DCM patients within the same US advanced heart failure program or between
any two such programs.

The prevalence of harboring a variant classified as P/LP among AA and EA probands with PPCM were 7.8% (95% CI, 0.0%-15.7%) and 29.5% (95% CI, 12.5%-46.5%), respectively (Table 3). The difference in prevalence by ancestry reflected substantially lower odds of having at least 1 P/LP for AA compared to EA probands with at least 1 P/LP/VUS variant (OR, 0.20; 95% CI, 0.09-0.44; Table 4). The prevalence of harboring a P/LP variant was similar in probands with PPCM and DCM within each ancestry group, reflecting similar odds of harboring P/LP/VUS variant noted above as well as similar odds of a P/LP variant among probands with at least 1 P/LP/VUS variant (OR, 1.12; 95% CI, 0.39-3.24; Table 4).

Using a simplified model with a variant set harmonized with a UK Biobank study^23^ (Supplemental Methods, “Comparison of Prevalence of Harboring P/LP Variants in PPCM Probands with UK Biobank”, eAppendix 1), the estimated prevalence of P/LP variants among the population of EA probands with PPCM at US advanced heart failure programs was 26.6% (95% CI, 12.6% - 40.6%) compared with 0.6% in the general population as represented in that study ^23^ (Supplemental Methods, eAppendix 1).

P/LP/VUS variants were classified depending on the PPCM classification of the proband(s) in which they were found and their characteristics compared (eTable 3). Variants found only in the PPCM group were comparable in terms of pathogenicity classification, gene evidence category, and predicted impact to those found only in the DCM group. Those found only in the PPCM group were similar to those found only in the DCM group in terms of interpretation criteria met, although the former had higher maximum alternate allele frequency in gnomAD non-founder populations (eTable 4).

### Familial aggregation in PPCM versus DCM

To understand familial aggregation of PPCM and DCM, clinical imaging data in FDRs were assessed to determine whether a DCM or pDCM phenotype was present. FDRs of AA probands had higher hazard of both DCM (Table e5) and DCM/pDCM (Table 5) than FDRs of EA probands after adjusting for FDR sex, proband enrollment site, proband ethnicity, proband PPCM status, and proband diagnosis age quartile. The hazard of DCM/pDCM was also higher for FDRs of probands diagnosed at younger ages (Table 5). Nonetheless, there was no evidence supporting reduced familial aggregation in FDRs of PPCM probands relative to those of female DCM probands for either DCM (hazard ratio, 0.58; 95% CI, 0.23 – 1.47; eTable 5) or DCM/pDCM (hazard ratio, 0.77; 95% CI, 0.47 – 1.28; Table 5, Figure 2b). For an FDR of a non-Hispanic EA proband with PPCM, the lowest estimated DCM/pDCM risk by age 80 was 26.8% (95% CI, 15.0%-45.0%) compared to 33.2% (95% CI, 21.2%-49.5%) for an FDR of a proband with DCM (Figure 2). For DCM risk by age 80, the corresponding estimates were 12.2% (95% CI, 4.5%-30.4%) and 20.0% (95% CI, 10.8%-35.3%). For a non-Hispanic EA proband with PPCM, DCM prevalence among FDRs in the lowest risk category aged 40-69 years estimated from this model (7.0% [95% CI, 0%-14.1%] females, 9.0% [95% CI, 1.6%-16.3%] males) was higher than general population estimates (0.30% females, 0.63% males) from a prior UK Biobank study.^27^

**Figure 2.**
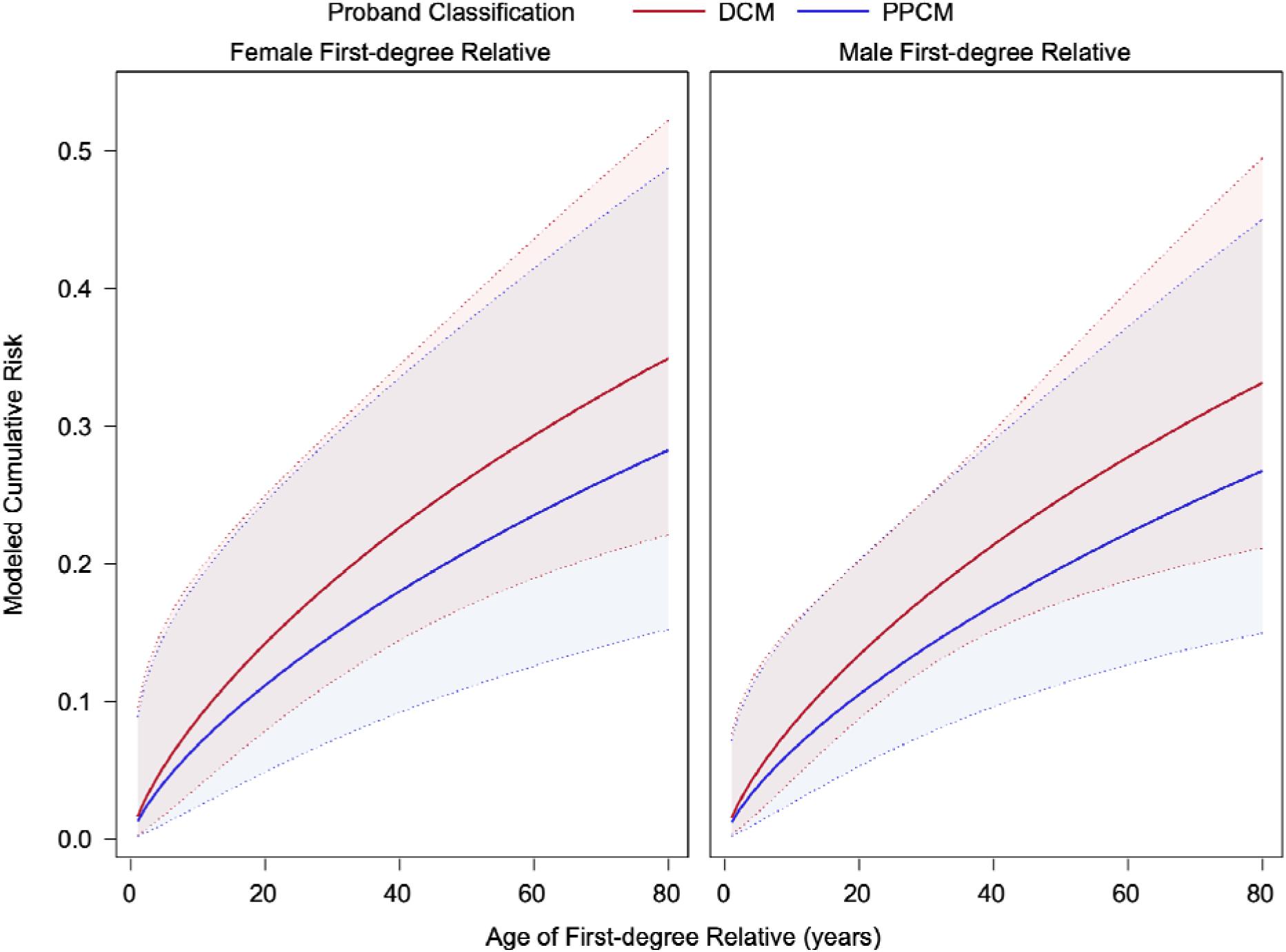
Model-based estimates of age-specific cumulative risk of DCM/partial DCM among the lowest-risk first-degree relatives of probands with either PPCM or DCM. A Weibull proportional hazards model for age-specific cumulative risk was fit to cross-sectional data on disease status at enrollment from first-degree relatives (Table 5). Age-specific cumulative risks and 95% confidence intervals of DCM/partial DCM are presented for female and male first-degree relatives of female Non-Hispanic European probands diagnosed with DCM or PPCM between ages 35.01 and 44.47 years seen at a typical US advanced heart failure program, defined as one at the mean or mode of the random effects distribution describing the population of such programs.

**Table 5.** Model fit for age-specific cumulative risk of DCM or partial DCM in a first-degree relative of a female proband with PPCM or DCM.

| Parameter<sup>a</sup> | Estimate | Standard Error | Hazard Ratio (95% CI) | P |
| --- | --- | --- | --- | --- |
| <u>Fixed Effects</u> |  |  |  |  |
| Weibull baseline survivor function parameters<sup>b</sup> |  |  |  |  |
| *a* | -4.0830 | 0.7915 | - | - |
| *b* | 0.7436 | 0.2286 | - | - |
| Proband PPCM classification |  |  |  |  |
| PPCM | -0.2575 | 0.2570 | 0.77 (0.47 - 1.28) | 0.32 |
| DCM | 0 | - | 1.00 | - |
| First-degree relative sex |  |  |  |  |
| Female | 0.06483 | 0.2365 | 1.07 (0.67 - 1.70) | 0.78 |
| Male | 0 | - | 1.00 | - |
| Proband ancestry group |  |  |  |  |
| African Ancestry | 0.3688 | 0.1680 | 1.45 (1.04 - 2.01) | 0.03 |
| European Ancestry | 0 | - | 1.00 | - |
| Proband self-reported ethnicity |  |  |  |  |
| Hispanic | -0.2249 | 0.3976 | 0.80 (0.37 – 1.74) | 0.57 |
| Non-Hispanic | 0 | - | 1.00 | - |
| Proband diagnosis age |  |  |  |  |
| I: [9.78, 34.99] | 0 | - | 1.00 | - |
| II: [35.01, 44.47] | -0.08501 | 0.3033 | 0.92 (0.51 – 1.66) | 0.78 |
| III: [44.48, 54.09] | -0.1198 | 0.3100 | 0.89 (0.48 – 1.63) | 0.70 |
| IV: [54.21, 78.31] | -0.5965 | 0.3070 | 0.55 (0.30 – 1.01) | 0.052 |
| <u>Variance Components</u> |  |  |  |  |
| Site ($\sigma^2_{\text{site}}$) | 0.03904 | 0.05139 | - | - |
| Residual | 0.9958 | 0.05543 | - | - |

CI = confidence interval; DCM = dilated cardiomyopathy; PPCM = peripartum cardiomyopathy.
<sup>a</sup> Parameters of a Weibull proportional hazards model for age-specific cumulative risk were estimated using a generalized estimating equation-type generalized linear mixed model with a binary outcome (presence or absence of DCM/partial DCM), complementary log-log link, random proband enrollment site intercept, and working independence correlation structure fit using residual subject specific pseudolikelihood. Bias-corrected robust standard errors were obtained using the Morel-Bokossa-Neerchal correction with sites as independent units. Two-sided p-values and Wald 95% confidence intervals were calculated using the standard normal distribution. Hazard ratios and their 95% confidence intervals were obtained by exponentiating corresponding estimates on the model
scale.
<sup>b</sup> $a$ and $b$ are parameters of the Weibull baseline survivor function of the form $S_o(t) = \exp[-\exp(a)t^b]$ . 665 first-degree relatives (103 of PPCM probands and 562 of DCM probands) contributed to this analysis.

## Discussion

The family-based design of the DCM Precision Medicine study offered a unique opportunity to assess the contribution of genetics to PPCM by evaluating the presence of shared genetic risk of DCM in FDRs of females with PPCM (Figure 3). Results from this analysis indicated that the estimated prevalence of DCM/pDCM in FDRs of PPCM probands was much higher than the general population and that the hazard of DCM/pDCM in FDRs of PPCM probands was not significantly lower than in FDRs of female probands with DCM (hazard ratio, 0.77; 95% CI, 0.47 – 1.28). In addition, the genetic findings in probands who had been pregnant were similar regardless of whether they were diagnosed with PPCM or DCM, confirming results previously found by others^5,7–9^. These complementary lines of evidence demonstrate a shared heritable genetic risk in PPCM similar to that which has been long established for DCM.

**Figure 3.**
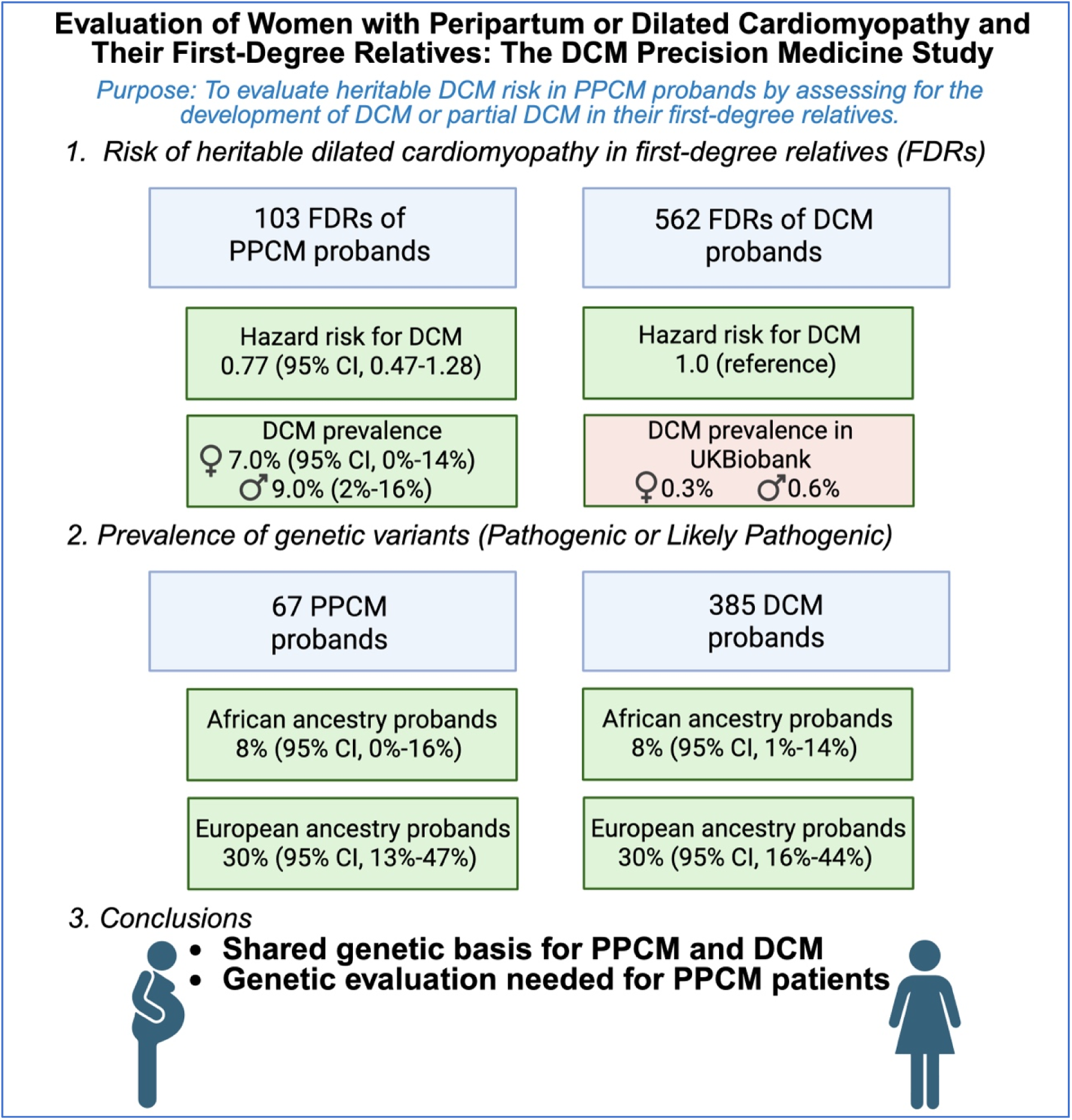
Central Figure. All probands (patients) in this study were females who had been pregnant so would have had similar risk of PPCM from pregnancy. The study showed, first, that the heritable risk of DCM was similar between first-degree relatives (FDRs) of women diagnosed with PPCM and women who were diagnosed with DCM, and that the DCM prevalence in both male and female FDRs was much higher than an estimate of DCM prevalence from the UKBiobank. Second, the study showed that the frequency of variants considered pathogenic and likely pathogenic for DCM were similar between probands with PPCM and DCM. In summary, FDR data from the DCM Precision Medicine Study provided a novel family-based analysis that showed a shared genetic basis of PPCM and DCM, all of which supports the need for genetic evaluations for all PPCM patients.

Although the genetic information identified in probands cannot be used for causal inference because probands were identified for study due to their DCM or PPCM phenotype, their FDRs were not identified based on a DCM or PPCM phenotype but only due to their relationship to the proband. This provided the opportunity to establish that similar genetic factors contributed to risk of PPCM and DCM in the probands by showing that DCM/pDCM risk was similar and higher than in the general population among their FDRs who share these genetic risk factors.

The utility of FDRs for assessing the impact of an environmental factor in a population at heightened genetic risk was also recently demonstrated in an analysis of the full DCM Precision Medicine Study cohort to assess the association between alcohol consumption and DCM. In that analysis, rare variant genetics were associated with prevalent DCM (odds ratio, 3.51, 95% CI: 2.33–5.29) in FDRs, but alcohol exposure was not independently associated, nor did it modify the association between rare variant genetics and DCM/pDCM in FDRs (P=0.55).^28^ As was the case with the analysis of DCM risk in FDRs related to alcohol use,^28^ this study was focused exclusively on assessing the underlying cause of PPCM/DCM and not on factors that modify the penetrance or expressivity of the putative genetic basis of PPCM. Such factors could include hypertension or other normal or aberrant pregnancy-related physiology.^1,2^ Such factors could trigger a genetic predisposition. Such factors, in association with the proband’s genetics, may also explain improvement or even resolution of DCM following the peripartum period. Also, PPCM does not universally have onset with a first pregnancy but can occur after one, two or more pregnancies, a fact that has defied explanation, but may be more fitting with genetics and its well-established age-dependent penetrance^29^ as an underlying causal factor.

The relatively small fraction, here less than one-quarter of probands, carrying P/LP variants might lead one to conclude that genetics are relevant for only a small fraction of PPCM or DCM patients. Three lines of evidence suggest a more expansive view of the contribution of genetics to both PPCM and DCM. First, based only on imaging-confirmed DCM/pDCM in FDRs in the whole DCM Precision Medicine Study cohort, an estimated 56.2% (Black) or 60.6% (White) of probands would have at least one living FDR meeting criteria for DCM/pDCM if all were screened with echocardiography.^11^ This level of familial recurrence strongly supports a genetic basis in more than half of probands. Second, the variant classification approach utilized for both the parent study^15^ and this study was selected to meet a highly stringent contemporary clinical standard in which the emphasis was on ensuring that a P/LP designation was not assigned without a very high level of evidence. While appropriate for use of variant for predictive (family-based) cascade testing, this requirement likely led to underestimation of rare variant genetic cause. Notably, nearly 45% of variants in the parent study were classified as VUS, twice the number of P/LP variants.^15^ While it is likely that not all of the variants classified as VUS were biologically relevant, it is also possible, if not likely, that many may have biological relevance. In particular, one-third of variants in the DCM Precision Medicine Study were novel (i.e., not present in any of the large population databases), and, unless those novel variants were truncating and thus much easier to classify as P/LP, nearly all were classified as a VUS due to limited evidence.^15^ Finally, this analysis has only focused on rare coding variants; data suggest that common variants are relevant for DCM risk,^30,31^ and low frequency variants and rare variants outside of coding regions may also contribute to DCM genetic cause.

Despite these data suggesting a more substantial genetic basis than reflected in the prevalence of P/LP variants, current data are insufficient to prove that all of DCM or PPCM has a genetic basis, which awaits larger confirmatory studies. Nevertheless, these findings are highly relevant for the clinical care because they suggest that *a genetic evaluation is indicated for all females who have PPCM*, which is still considered elective by many cardiologists, or only indicated with a compelling family history of DCM. A genetic evaluation includes a careful family history for cardiovascular disease and genetic testing with pre- and post-test counseling.^32^ The knowledge derived from such a genetic evaluation will directly contribute to the care of the patient’s cardiomyopathy and may also inform her considerations for possible future pregnancies. Genetic information also can contribute to family-based care, including risk assessments for heritable cardiomyopathy in FDRs,^32^ including any children of a PPCM patient.

The findings of this study are also highly relevant for public health, as PPCM is a leading cause of maternal mortality,^3^ and maternal mortality driven in large part by PPCM is higher in Black females for a variety of reasons beyond genetics, including social determinants of health.^3,33–35^ The evidence provided here that DCM genetics are also key to assessing PPCM risk is relevant for all females with PPCM, but particularly so for females of AA due to disparities in access, genetic testing utilization, and representation in genetic databases for non-European individuals.^36,37^ Complicating genetic assessments of PPCM and DCM for individuals of AA is the currently incomplete knowledge of the differences in the genetic architecture of DCM in AA versus EA,^15^ as the fraction and number of P/LP variants, particularly of *TTN* truncating variants, were lower in probands of AA compared to EA.^15^ While the data from this study did not show increased risk of PPCM in females of AA compared to females of EA, prior data suggest that DCM risk more generally may be increased in individuals of AA relative to individuals of EA. Our findings of increased age-specific cumulative risks of DCM (HR 2.18, 95% CI 1.39 - 3.43, p = 0.001) and DCM/partial DCM (HR of 1.45; 95% CI 1.04 – 2.01; p = 0.03) among FDRs of AA female probands shown in this study support this possibility. An earlier case-control study also observed an increased risk in both male and female patients of AA with DCM.^38^ Our prior family-based observations also support a possible increased risk of DCM in AA, where the estimated prevalence of familial DCM was higher in Black probands than in White probands (difference 11.3%, 95% CI, 1.9% - 20.8%) and the model-based estimates of age-specific cumulative risk were higher in FDRs of Black probands compared to those of White probands.^11^

Importantly, only female probands who had been pregnant at least once were included in the current analysis so that all probands would have been exposed to any risk arising from pregnancy. Our observation of similar frequencies of P/LP variants in patients with either PPCM or DCM has been previously reported,^7,8^ which also supports the conclusion that genetics likely underlies PPCM. The availability of both PPCM and DCM groups enrolled with the same inclusion/exclusion criteria is a strength of our study, whereas previous studies of PPCM have been less able to directly compare PPCM to DCM.

## Limitations

Large confidence intervals for this study were related to the limited numbers of PPCM patients; a much larger family-based cohort of systematically enrolled females with PPCM and DCM is needed to confirm and extend these observations. Long-term outcomes data would also provide additional information regarding genetic risk. Also, females with PPCM but without LV dilation would have been excluded, as LV dilation was an inclusion criterion for all probands, and a smaller LV diameter has been associated with increased likelihood of recovery of function.^39^ Also, since probands were recruited from advanced heart failure clinics, our results may not be generalizable to all PPCM patients. Nevertheless, the PPCM patients reported here represent a cohort with advanced disease and undoubtedly an outsized contribution to maternal morbidity and mortality, providing relevant findings for the field.

## Conclusion

The risks of DCM or pDCM in FDRs of females with PPCM were similar to those in FDRs of females with DCM, and the prevalence of DCM in their FDRs was much higher than the general population. Also, the prevalence of P/LP/VUS and P/LP findings among females with PPCM were not different from females with DCM, and the prevalence of P/LP findings in EA probands was far higher than in the general population. These results provide complementary lines of evidence supporting a shared genetic basis for PPCM and DCM. This study suggests that genetic evaluations are indicated for all females with PPCM.

## Supporting information

Kransdorf et al Supplemental Methods and Data

## Data Availability

The data are available at dbGaP; additional data can be made available with reasonable request.

## Funding

Research reported in this publication was supported by a parent award from the National Heart, Lung, And Blood Institute of the National Institutes of Health under Award Number R01HL128857 to Dr. Hershberger, which included a supplement from the National Human Genome Research Institute. The content is solely the responsibility of the authors and does not necessarily represent the official views of the National Institutes of Health. Support was also provided to the DCM Research Project from the Graciano Family Fund.

## Disclosures

The authors declare no conflicts or competing interests.

## Acknowledgements

The investigators thank the families with DCM who have participated in this study, without whom this effort would not be possible. The DCM Precision Medicine Study was supported by computational infrastructure provided by The Ohio State University Division of Human Genetics Data Management Platform and the Ohio Supercomputer Center.

## Abbreviations List

FDR/FDRs: first-degree relative, first-degree relatives
LP: likely pathogenic
P: pathogenic
pDCM: partial DCM
PPCM: peripartum cardiomyopathy
VUS: variant of uncertain significance

## Notes

### Competing Interest Statement

The authors have declared no competing interest.

### Author Declarations

The Institutional Review Boards (IRB) at the Ohio State University and all clinical sites approved the initial study, followed by single IRB oversight at the University of Pennsylvania. All participants gave written informed consent.

### Summary of Updates

This is the first revised submission.

