## Supplementary material for "Evaluation of Women with Peripartum or Dilated Cardiomyopathy and Their First-Degree Relatives: The DCM Precision Medicine Study": Kransdorf et al Supplemental Methods and Data


**eAppendix 1. Supplemental Methods**

**eTable 1. Model fit for the probability of PPCM in the population of female probands with specified characteristics at a particular US advanced heart failure program**

**eTable 2. Variants classified as P, LP, or VUS, by gene and PPCM classification of female proband(s) in which they were observed**

**eTable 3. Characteristics of variants classified as P, LP, or VUS observed in female probands by PPCM classification**

**eTable 4. Post-hoc pairwise comparisons of variant characteristics that differed across PPCM classifications with P<0.05**

**eTable 5. Model fit for the age-specific cumulative risk of DCM only in a first-degree relative of a female proband with PPCM or DCM**

**eReferences**

### **eAppendix 1.** Supplemental Methods

**Statistical Methods**

Throughout, let $k = 1, \ldots, 28$ index the proband's enrollment site, $j = 1, \ldots, n_k$ index the probands at site $k$, and $i = 1, \ldots, n_{jk}$ index the included first-degree relatives of proband $jk$. Twenty-six enrollment sites were advanced heart failure programs (one was inactivated after 3 probands were enrolled leaving 25 who completed the study), one was a geographically remote satellite site of a such a program, and one was a virtual site at the coordinating center. This study included an embedded open-label randomized controlled trial of a behavioral intervention that was administered to probands in order to increase screening and surveillance uptake in family members.[1] Analyses described below included families in both the control and intervention arms because the analysis would be unaffected by variation in screening uptake among families as long as other assumptions were correct.

Generalized Cochran-Mantel-Haenszel Test with Randomly Selected Sites

The test statistic $T_{EL} = G' V_G^{-1} G$ was used with proband enrollment sites as primary sampling units, which treated site effects as random in a manner consistent with our more complex models.[2,3] The statistic $G$ was determined by the row and column scoring appropriate for a general association hypothesis, and $V_G$ was its sample covariance matrix across sites. Two-sided p-values were calculated by comparing $(q - df)T_{EL}/\big(df(q-1)\big)$ to a $F(df, q - df)$ distribution, where $q$ is the number of sites and $df = (R-1)(C-1)$ for a table with $R$ rows and $C$ columns. The p-value is designed to detect within-site differences in the distribution of the column variable across rows that are consistent across sites.

Boos-Brownie Test with Randomly Selected Sites

The relative effect for site $k$, $\theta_k$, is the probability that a randomly selected value from the peripartum cardiomyopathy (PPCM) group was greater than a randomly selected value from the dilated cardiomyopathy (DCM) group.[4,5] If a variable has no tendency to take larger or smaller values in PPCM patients compared to DCM patients at a particular site, then $\theta_k = 1/2$ . The $\theta_k$ for each informative site with at least one proband in each group was estimated from the within-site midranks of the continuous variable, and the test statistic $V_1$, which is a one-sample $t$ statistic for the null that the mean of these estimates across sites equals 1/2, was used to test the null hypothesis that $\theta_k = 1/2$ for all $k$.[4] Like $T_{EL}$, $V_1$ treats site effects as being random and is asymptotically normally distributed as the number of sites $q$ grows large.[4] Two-sided p-values were calculated by comparing $V_1$ to a $t$ distribution with $q - 1$ degrees of freedom. The p-value is meant to detect variables that tend to take larger or smaller values in PPCM patients compared to DCM patients consistently at each site.

Logit Model for Ancestry-Specific Prevalence of PPCM Among Female Probands

For proband $j$ at site $k$, let $l_{jk} = 1$ (European), 2 (African) denote the ancestry group, $m_{jk} = 1$ (Non-Hispanic), 2 (Hispanic) denote the ethnicity, $r_{jk} = 1, 2, 3, 4$ denote the enrollment age quartile, and $Y_{jk}$ denote the binary outcome equaling 1 if the proband had PPCM and 0 otherwise.

The target of inference was the probability of PPCM in an eligible proband conditional on site and other variables included in the model. A generalized linear mixed model (GLMM)[6] for the binary disease classification was defined using the logit link as follows:

$$\pi_{jk} = \text{logistic}\left(\mu + \alpha_{l_{jk}} + \beta_{m_{jk}} + \gamma_{r_{jk}} + a_k\right) \tag{1}$$

where $\mu$, $\alpha_{l_{jk}}$, $\beta_{m_{jk}}$, and $\gamma_{r_{jk}}$ are fixed effects with $\alpha_1 \equiv \beta_1 \equiv \gamma_4 \equiv 0$, and $a_k \sim N(0, \sigma^2_{\text{site}})$ are random effects independently distributed across sites.

Under this model, $\exp(\alpha_l)$ is the ratio of the odds of having PPCM in a proband in ancestry group $l$ to those in a comparable proband in the European ancestry group ($l = 1$). The odds ratios $\exp(\beta_m)$ and $\exp(\gamma_r)$ are interpreted similarly. If $\sigma^2_{\text{site}} > 0$, all odds ratios can be interpreted as comparing probands within the same US advanced heart failure program. However, if $\sigma^2_{\text{site}} = 0$, these odds ratios can also be interpreted as comparing probands between any two such programs.

Parameters of the model in (1) were estimated using residual subject-specific pseudolikelihood (RSPL)[6-8] as implemented in SAS/STAT PROC GLIMMIX. This approach is based on fitting an approximating linear mixed model to a transformed pseudo-response and works well for nearly continuous outcomes or discrete outcomes when there are a large number of observations per independent unit (i.e., site).[6-9] Standard linear mixed model inferential procedures can then be applied to the resulting fixed effects estimates, which have an approximate multivariate normal distribution centered at the true parameter values.[6,7] Inference on the model effects was performed using the Morel-Bokossa-Neerchal (MBN) bias-corrected estimate of the empirical covariance matrix and the standard normal distribution.[7,10] Odds ratios as well as their Wald 95% confidence intervals (CIs) and two-sided Wald p-values for the null hypothesis that the odds ratio was 1 are shown in eTable 1. This model was also used to obtain ancestry-specific marginally standardized estimates of outcome probabilities for probands seen at US advanced heart failure programs, as described below.

Hierarchical Logit Models for Trinomial Outcomes in Probands

Our previously published model[11] for trinomial outcomes was adapted for the current analysis; its development is recapitulated below with modifications specific to the current analysis. For proband $j$ at site $k$, let $l_{jk} =$ 1 (European), 2 (African) denote the ancestry group, $m_{jk} =$ 1 (Non-Hispanic), 2 (Hispanic) denote the ethnicity, $r_{jk} =$ 1, 2, 3, 4 denote the DCM diagnosis age quartile, $s_{jk} =$ 1 (DCM), 2 (PPCM) denote the disease classification, and $\mathbf{Y}_{jk} = \left(Y_{jk1}, Y_{jk2}, Y_{jk3} = 1 - Y_{jk1} - Y_{jk2}\right)$ be a vector of indicator variables, where $Y_{jkc} = 1$ and $Y_{jkc'} = 0$ for $c' \neq c$ if proband $jk$ is in category $c$ of three possible mutually exclusive and exhaustive categories. Conditional on proband enrollment site and covariates, this random vector is a single trial from a multinomial distribution with parameters $\left(\pi_{jk1}, \pi_{jk2}, 1 - \pi_{jk1} - \pi_{jk2}\right)$. Using the well-known factorization of the multinomial probability into binomial conditional probabilities along with reparameterization yields:[12]

$$\begin{aligned}\Pr\left(\mathbf{Y}_{jk}\right) &= \pi_{ij1}^{Y_{ij1}}\left(1 - \pi_{jk1}\right)^{1-Y_{jk1}}\left(\frac{\pi_{jk2}}{1 - \pi_{jk1}}\right)^{Y_{jk2}}\left(\frac{1 - \pi_{jk1} - \pi_{jk2}}{1 - \pi_{jk1}}\right)^{1-Y_{jk1}-Y_{jk2}} \\ &= p_{jk1}^{Y_{jk1}}\left(1 - p_{jk1}\right)^{1-Y_{jk1}} p_{jk2}^{Y_{jk2}}\left(1 - p_{jk2}\right)^{1-Y_{jk1}-Y_{jk2}} \\ &= \begin{cases} p_{jk1} & \text{if } \mathbf{Y}_{jk} = (1,0,0) \\ \left(1 - p_{jk1}\right)p_{jk2} & \text{if } \mathbf{Y}_{jk} = (0,1,0) \\ \left(1 - p_{jk1}\right)\left(1 - p_{jk2}\right) & \text{if } \mathbf{Y}_{jk} = (0,0,1) \end{cases}\end{aligned} \tag{2}$$

where $p_{jk1} = \pi_{jk1}$, the marginal probability of falling in category 1, and $p_{jk2} = \pi_{jk2}/\left(1 - \pi_{jk1}\right)$, the probability of falling in category 2 conditional on not falling in category 1 (or conditional on falling in categories 2 or 3).

The probabilities $p_{jk1}$ and $p_{jk2}$ for proband $j$ at site $k$ were modeled as a function of disease classification, ancestry group, ethnicity, and DCM diagnosis age quartile fixed effects as well as a random effect to account for site heterogeneity using the logit link:

$$\operatorname{logit}\left(p_{jkg}\right) = \zeta_g + \alpha_{l_{jkg}} + \beta_{m_{jkg}} + \gamma_{r_{jkg}} + \delta_{s_{jkg}} + u_{kg} \tag{3}$$

where $\zeta_g$, $\alpha_{l_{jkg}}$, $\beta_{m_{jkg}}$, $\gamma_{r_{jkg}}$, and $\delta_{s_{jkg}}$ are fixed effects with $\alpha_{1g} \equiv \beta_{1g} \equiv \gamma_{1g} \equiv \delta_{1g} \equiv 0$ for $g = 1,2$ and $\mathbf{u}_{\boldsymbol{k}} = \begin{bmatrix} u_{k1} \\ u_{k2} \end{bmatrix} \sim N\left(0, \begin{bmatrix} \sigma_1^2 & \sigma_{12} \\ \sigma_{12} & \sigma_2^2 \end{bmatrix}\right)$ is a vector of random effects with an unstructured covariance matrix independently distributed across sites.

Equations (2) and (3) define a GLMM[6,7] in which the marginal likelihood for each independent unit (i.e., site) with $n_k$ probands is given by:

$$L_k(\boldsymbol{\theta}, \boldsymbol{\sigma}) = \iint_{-\infty}^{+\infty} \prod_{i=1}^{n_k} \left( \begin{array}{c} p_{jk1}^{I[\mathbf{Y}_{jk}=(1,0,0)]}\left[\left(1 - p_{jk1}\right)p_{jk2}\right]^{I[\mathbf{Y}_{jk}=(0,1,0)]} \\ \times \left[\left(1 - p_{jk1}\right)\left(1 - p_{jk2}\right)\right]^{I[\mathbf{Y}_{jk}=(0,0,1)]} \end{array} \right) \phi(u_{k1}, u_{k2}, \boldsymbol{\sigma}) du_{k1} du_{k2} \tag{4}$$

where $\boldsymbol{\theta}$ denotes the fixed effects parameters in $p_{jkg}$, $\boldsymbol{\sigma}$ denotes the variance components, and $\phi$ is the multivariate normal density. Note that this likelihood component is identical to one that would be obtained if a proband with $\mathbf{Y}_{jk} =$ (1,0,0) had one observation that was a success for a binary random variable with success probability $p_{jk1}$; a proband with $\mathbf{Y}_{jk} = (0,1,0)$ had two observations, a failure and a success, for conditionally independent binary random variables with respective success probabilities $p_{jk1}$ and $p_{jk2}$; and a proband with $\mathbf{Y}_{ij} = (0,0,1)$ had two observations, both failures, for conditionally independent binary random variables with respective success probabilities $p_{jk1}$ and

$p_{jk2}$. The marginal likelihood comprising the product of components in (4) across sites can therefore be maximized using standard GLMM software applied to a binary outcome with one or two observations per proband obtained as defined above.

Under this model, $\exp(\alpha_{l1})$ is the ratio of the odds having an outcome in category 1 in a proband in ancestry group $l$ to those in a comparable proband in the European ancestry group ($l = 1$). Likewise, $\exp(\alpha_{l2})$ is the ratio of the odds of having an outcome in category 2 in a proband in ancestry group $l$ who did not have an outcome in category 1 to those of a comparable proband in the European ancestry group. The odds ratios $\exp(\beta_{m1})$, $\exp(\beta_{m2})$, $\exp(\gamma_{r1})$, $\exp(\gamma_{r2})$, $\exp(\delta_{s1})$, and $\exp(\delta_{s2})$ are interpreted similarly. If $\sigma_k^2 > 0$, all odds ratios for that $k$ can be interpreted as comparing probands within the same US advanced heart failure program. However, if $\sigma_k^2 = 0$, these odds ratios can also be interpreted as comparing probands between any two such programs.

Parameters of the model in (3) and (4) were initially estimated using RSPL[6-8] as implemented in SAS/STAT PROC GLIMMIX. Due to the Cauchy-Schwartz inequality, $\sigma_{12} = 0$ unless both $\sigma_1^2$ and $\sigma_2^2$ are non-zero, so initial model fits used a variance components structure in which $\sigma_{12} = 0$ to determine if there was any evidence of site heterogeneity for either or both outcomes. The model fits for both outcomes yielded estimates of zero for $\sigma_2^2$ only, so the models were refit using RSPL including $u_{k1}$ only. Odds ratios as well as their Wald 95% CIs and two-sided Wald p-values for the null hypothesis that the odds ratio was 1 are shown in Table 4. All p-values and CIs were produced using the MBN bias-corrected estimate of the empirical covariance matrix and the standard normal distribution.[7,10] This model was also used to obtain marginally standardized estimates of outcome probabilities for probands with a particular disease classification seen at US advanced heart failure programs, as described below.

Comparison of Prevalence of Harboring P/LP Variants in PPCM Probands with UK Biobank

The prevalence of harboring P/LP variants in a common set of genes was compared between PPCM probands from the DCM Precision Medicine Study and the general population of European ancestry based on the data provided by Shah and colleagues.[13] These authors applied several filtering strategies to identify putatively pathogenic rare variants in a set of DCM genes among 18665 individuals (96.8% White) from the UK Biobank. The one most similar to the one used by the DCM Precision Medicine Study was the InterVAR FAF strategy, which used ACMG-based variant interpretation and a filtering allele frequency of $\leq 8.4 \times 10^{-5}$. To obtain the best comparison possible between the two studies, we considered variants only in genes that were also included in the DCM Precision Medicine Study, which eliminated 40 individuals harboring P/LP variants in *CTF1*, *DTNA*, *GATAD1*, *MYL2*, NKX2-5, *OBSCN*, *PLEKHM2*, *PRDM16*, *PSEN2*, *TBX20*, and *TNNI3K* (see their Table S8). The estimated prevalence of harboring P/LP variants in the European ancestry general population obtained from this study was therefore (154 – 40)/18665 = 0.6%.

For the DCM Precision Medicine Study, we eliminated variants in three genes not considered by Shah and colleagues, which included *CRYAB*, *PDLIM3*, and *PKP2* (none P/LP). We also eliminated 1 variant classified as P/LP in the DCM Precision Medicine Study with alternate allele frequency $>8.4 \times 10^{-5}$ in at least one gnomAD v2.1.1 non-founder population.[14] A GLMM for binary carrier status of proband $j$ at site $k$ for the remaining P/LP variants was defined using the logit link as follows:

$$\operatorname{logit}(\pi_{jk}) = \mu + \alpha_{l_{jk}} + \beta_{m_{jk}} + \gamma_{r_{jk}} + \delta_{s_{jk}} + u_k \quad (5)$$

where $l_{jk} = 1$ (European), 2 (African) denotes the ancestry group, $m_{jk} = 1$ (Non-Hispanic), 2 (Hispanic) denotes the ethnicity, $r_{jk} = 1, 2, 3, 4$ denotes the DCM diagnosis age quartile, $s_{jk} = 1$ (DCM), 2 (PPCM) denotes the disease classification, and $\mu$, $\alpha_{l_{jk}}$, $\beta_{m_{jk}}$, $\gamma_{r_{jk}}$, and $\delta_{s_{jk}}$ are fixed effects with $\alpha_1 \equiv \beta_1 \equiv \gamma_1 \equiv \delta_1 \equiv 0$, and $u_k \sim N(0, \sigma_{\text{site}}^2)$ are random effects independently distributed across sites.

Parameters of the model in (5) were initially estimated using RSPL[6-8] as implemented in SAS/STAT PROC GLIMMIX. This model fit yielded an estimate of zero for $\sigma_{\text{site}}^2$, in which case the model reduces to a standard generalized linear model for a binary outcome with independent observations and RSPL is equivalent to maximum likelihood. Inference on the model effects was therefore performed in a maximum likelihood framework using the inverse of the observed information matrix to estimate the covariance matrix of the fixed effects. Fixed effects estimates and their covariance matrix were used to obtain marginally standardized estimates of the prevalence of harboring P/LP variants among PPCM probands seen at US advanced heart failure programs, as described below.

Marginally Standardized Estimates

In addition to identifying factors associated with the outcome, the models in (1) and (3) can produce two types of estimates: conditional estimates for a single advanced heart failure program in the US or marginal estimates across all advanced heart failure programs in the US. These two types of estimates will not be equal unless there is no heterogeneity in these programs,[7,8,15] and the appropriate choice depends on how the estimates will be applied.[16,17] In the current context, we expect that these estimates will be used by clinicians to understand risks among different groups at their programs as well as for comparison with prior single-center studies, in which case conditional estimates for a single advanced heart failure program in the US are most relevant.[16,17] For advanced heart failure programs in our study, empirical Bayes predictions of the random effects $a_k$ and $\mathbf{u}_k$ could have been used to generate program-specific estimates,[7,16-18] but these would not apply to an external program not included in our sample.[7,16,17] To facilitate application at such a program, conditional estimates for a typical advanced heart failure program in the US with $a_k = 0$ (or $\mathbf{u}_k = 0$),[7,8,15] which have the best expected within-program calibration for external programs,[16,17] were presented. Such a typical US advanced heart failure program is defined by being at the mean or mode of the random effects distribution describing the population of such programs in the US.[7,8,15-17]

To generate marginally standardized[19] ancestry-specific PPCM prevalence estimates at a typical US advanced heart failure program, the weighted average of probabilities derived from (1) was taken over the distribution of $\{m, r\}$ in each ancestry group assuming balance across the four possible enrollment age quartiles and a proportion of Hispanic probands equal to the 2021 US census population estimates (18.9%).[19,20] The delta method[21] as implemented in SAS/STAT PROC NLMIXED was applied to the parameter estimates for the model in (1) and their estimated covariance matrix to obtain estimates and standard errors for the marginally standardized probabilities. The resulting estimates and 95% CIs are reported in the manuscript text.

Because the dependence of $p_{jkg}$ on $j$ and $k$ in the hierarchical logit model in (3) and (4) derives exclusively from $\{z_{jk}, u_{kg}\}$, it can be expressed as a function of these quantities, $p_{jkg} = p_g(z_{jk}, u_{kg})$. As the estimated variance component for $u_{k2}$ was zero for both outcomes, $p_{jk2} = p_2(z_{jk})$ in the final model fits in which the random effect was omitted for this linear predictor. Letting $z = \{l, m, r, s\}$ denote a particular possible value of $z_{jk} = \{l_{jk}, m_{jk}, r_{jk}, s_{jk}\}$, the multinomial probabilities in the subpopulation of probands with characteristics $z$ at a typical US advanced heart failure program were obtained as:

$$\Pr(\mathbf{Y}_{jk} | z, u_{k1} = 0) = \begin{cases} p_1(z, 0) & \text{if } \mathbf{Y}_{jk} = (1,0,0) \\ (1 - p_1(z, 0))p_2(z) & \text{if } \mathbf{Y}_{jk} = (0,1,0) \\ (1 - p_1(z, 0))(1 - p_2(z)) & \text{if } \mathbf{Y}_{jk} = (0,0,1) \end{cases} \quad (6)$$

To generate marginally standardized ancestry- and disease-specific probabilities that apply to a proband seen at a typical US advanced heart failure program, the weighted average of each multinomial probability in (6) was taken over the distribution of $\{m, r\}$ in each combination of ancestry group and disease classification assuming balance across the two youngest DCM diagnosis age quartiles and a proportion of Hispanic probands equal to the 2021 US census population estimates (18.9%).[19,20] Only the two youngest DCM diagnosis age quartiles were considered because no probands with PPCM were observed in the older age quartiles encompassing ages ≥44.48 years. The delta method[21] as implemented in SAS/STAT PROC NLMIXED was applied to the parameter estimates for the model in (3) and (4) and their estimated covariance matrix to obtain estimates and standard errors for the marginally standardized multinomial probabilities. Joint Wald 95% CIs with simultaneous 95% coverage over all 3 multinomial probabilities were obtained using standard normal quantiles and the Bonferroni correction. The resulting estimates and joint 95% CIs are presented in Table 3.

Finally, for comparison with population P/LP carrier prevalence estimates from the Shah and colleagues UK Biobank study, a marginal estimate from the model in (5) applying to the entire population of PPCM probands of European ancestry seen at US advanced heart failure programs was needed. Because the final model fit in (5) did not include $u_k$ due to its zero estimated variance component, these could be obtained directly as predicted probabilities from the model. The weighted average of these probabilities was taken over the distribution of $\{m, r\}$ in PPCM probands of European ancestry assuming balance across the two youngest DCM diagnosis age quartiles and a proportion of Hispanic probands equal to the 2021 US census population estimates (18.9%).[19,20] The delta method[21] as implemented in SAS/STAT PROC NLMIXED was applied to the parameter estimates for the model in (5) and their estimated covariance matrix to obtain estimates and standard errors for the marginally standardized probability. The resulting estimate and its 95% CI are reported in the manuscript text.

Age-Specific Cumulative Risk for First-Degree Relatives

Our previously published model[22] for estimating the age-specific cumulative risk of DCM and partial phenotypes was adapted for the current analysis; its development is recapitulated below with modifications specific to the current analysis. For each screened first-degree relative, it was possible to determine whether DCM or partial phenotypes were present by the age of enrollment. This observation scheme yielded current status data, a special type of interval-censored survival data[23-25] that can be used to estimate age-specific cumulative risks. Let $i = 1, \ldots, n_{jk}$ index the screened first-degree relatives of proband $jk$, $l_{jk} = 1$ (European), 2 (African) denote the proband's ancestry group, $m_{jk} = 1$ (Non-Hispanic), 2 (Hispanic) denote the proband's ethnicity, $r_{jk} = 1, 2, 3, 4$ denote the proband's DCM diagnosis age quartile, $s_{jk} = 1$ (DCM), 2 (PPCM) denote the proband's disease classification, and $w_{ijk} = 1$ (male), 2 (female) denote first-degree relative sex. Age at disease onset, $T_{ijk}$, in a first-degree relative with characteristics $\{l_{jk}, m_{jk}, r_{jk}, s_{jk}, w_{ijk}\}$ at site $k$ was assumed to have a marginal distribution with a Weibull baseline survivor function $S_0(t) = \exp[-\exp(a)\, t^b]$ influenced by covariates and random effects through a proportional hazards model,[23-25] yielding the following marginal survivor function:

$$S\big(t \big| l_{jk}, m_{jk}, r_{jk}, s_{jk}, w_{ijk}, u_k\big) = \exp\left[-\exp(a)\, t^b \exp\left(\alpha_{l_{jk}} + \beta_{m_{jk}} + \gamma_{r_{jk}} + \delta_{s_{jk}} + \omega_{w_{ijk}} + u_k\right)\right] \tag{7}$$

where $\alpha_{l_{jk}}$, $\beta_{m_{jk}}$, $\gamma_{r_{jk}}$, $\delta_{s_{jk}}$, and $\omega_{w_{ijk}}$ are fixed effects with $\alpha_1 \equiv \beta_1 \equiv \gamma_1 \equiv \delta_1 \equiv \omega_1 \equiv 0$ and $u_k \sim N(0, \sigma^2_{\text{site}})$ are random effects designed to reflect site-specific random variation in the marginal disease hazard for a first-degree relative. Under this model, $\exp(\delta_s)$ is the ratio of the disease hazard for a first-degree relative of a proband with disease classification $s$ to that for a first-degree relative of a proband with a DCM disease classification at the same site who is identical on all other factors. If $\sigma^2_{\text{site}} = 0$, this hazard ratio can also be interpreted as comparing first-degree relatives between any two sites. The hazard ratios $\exp(\alpha_l)$, $\exp(\beta_m)$, $\exp(\gamma_r)$, and $\exp(\omega_w)$ are interpreted similarly.

The parameters in (7) were estimated as follows using current status data. The age at disease onset was unobserved because DCM and partial phenotypes are typically asymptomatic for months or years before presentation. However, enrolling a first-degree relative at a particular age ($C_{ijk}$) and examining him or her allowed for determination of whether $T_{ijk}$ was before or after $C_{ijk}$ on the basis of whether the individual had disease at $C_{ijk}$. Defining the observable random variable $Y_{ijk} = I\big(T_{ijk} \le c_{ijk}\big)$ and assuming conditional independence of $C_{ijk}$ and $T_{ijk}$ given $\{l_{jk}, m_{jk}, r_{jk}, s_{jk}, w_{ijk}, u_k\}$ yielded:

$$\begin{aligned} &\Pr\big(Y_{ijk} \big| C_{ijk} = c_{ijk}, l_{jk}, m_{jk}, r_{jk}, s_{jk}, w_{ijk}, u_k\big) \\ &= \Pr\big(Y_{ijk} \big| l_{jk}, m_{jk}, r_{jk}, s_{jk}, w_{ijk}, u_k\big) \\ &= \big[1 - S\big(c_{ijk} \big| l_{jk}, m_{jk}, r_{jk}, s_{jk}, w_{ijk}, u_k\big)\big]^{Y_{ijk}} S\big(c_{ijk} \big| l_{jk}, m_{jk}, r_{jk}, s_{jk}, w_{ijk}, u_k\big)^{1-Y_{ijk}} \end{aligned} \tag{8}$$

Thus, $Y_{ijk}$ was a Bernoulli random variable with conditional success probability $\Pr\big(Y_{ijk} = 1 \big| c_{ijk}, l_{jk}, m_{jk}, r_{jk}, s_{jk}, w_{ijk}, u_k\big) = 1 - S\big(c_{ijk} \big| l_{jk}, m_{jk}, r_{jk}, s_{jk}, w_{ijk}, u_k\big)$. Furthermore, this probability can be related to the parameters in (7) by applying the complementary log-log link:

$$\begin{aligned} &\ln\big(-\ln\big(1 - \Pr\big(Y_{ir} = 1 | c_{ijk}, l_{jk}, m_{jk}, r_{jk}, s_{jk}, w_{ijk}, u_k\big)\big)\big) \\ &= a + b \ln c_{ijk} + \alpha_{l_{jk}} + \beta_{m_{jk}} + \gamma_{r_{jk}} + \delta_{s_{jk}} + \omega_{w_{ijk}} + u_k \end{aligned} \tag{9}$$

With a single first-degree relative per proband, the model could have been fit using a standard GLMM with a binary outcome, site-level random effects, a complementary log-log link, and a linear predictor given by (9). However, with multiple first-degree relatives per proband, the effect of non-independence on estimated standard errors needed to be taken into account. The parameters and variance components for the model in (9) were therefore estimated using a moments-based or generalized estimating equation (GEE)-type GLMM[6,7] with a binary outcome, site-level random effects, a complementary log-log link, a linear predictor given by (9), and a working independence correlation matrix for first-degree relatives within each family. This model was fit with RSPL as implemented in SAS/STAT PROC GLIMMIX; if the model fit yielded an estimate of zero for $\sigma^2_{\text{site}}$, the model with $u_k$ omitted was refit using RSPL. In either case, inference on fixed effects used the MBN corrected empirical covariance estimator with sites as independent units to account for residual within-family correlation.[6,7,10] Estimation within this framework assumed that, given covariates and site, first-degree relative participation did not depend on $Y_{ijk}$.

DCM or partial phenotypes in a first-degree relative were required to have no known cause for $Y_{ijk} = 1$. Thus, $Y_{ijk} = 0$ for individuals without left ventricular systolic dysfunction or left ventricular enlargement as well as for those with DCM or partial phenotypes with a probable environmental cause. This approach is tantamount to assuming that first-degree relatives with a DCM phenotype arising from an environmental cause would not have developed disease absent this exposure. As these first-degree relatives were genetically at-risk, they may still have developed disease absent the exposure, and so this approach likely underestimates the age-specific cumulative risk.

Parameter estimates, standard errors, and hazard ratios with Wald 95% CIs constructed using standard normal quantiles are presented in Table 5 and eTable 5. The age-specific cumulative risk for a first-degree relative with characteristics $\{l, m, r, s, w, u_k\}$ at a typical US advanced heart failure program was obtained as:

$$F(t|l, m, r, s, w, u_k = 0) = 1 - \exp[-\exp(a)\, t^b \exp(\alpha_l + \beta_m + \gamma_r + \delta_s + \omega_w)] \quad (10)$$

The value of $\{l, m, r, s, w\}$ was chosen to reflect the lowest-risk first-degree relatives of a Non-Hispanic European proband in the youngest two DCM age quartiles, as in the implied DCM prevalence analysis below. The delta method[21] as implemented in SAS/STAT PROC NLMIXED was applied to the parameter estimates for the model in (9) and their estimated covariance matrix to obtain estimates and standard errors for the probabilities in (10) at a given age. The resulting estimates and pointwise 95% CIs are shown in Figure 1.

Implied DCM Prevalence Among First-Degree Relatives

DCM prevalence among first-degree relatives of PPCM probands of non-Hispanic European ancestry aged 40 to 69 was estimated from the model in (9) for comparison with imaging-based estimates of population DCM prevalence from the UK Biobank.[26] Because $\sigma^2_{\text{site}} = 0$ for DCM, the marginal age-specific cumulative risk of DCM at age $t$ could be obtained directly from (10). A lower bound on the implied prevalence among the lowest-risk group of these first-degree relatives of sex $w$ between 40 and 69 years of age was then given by:

$$\begin{aligned}
&\int_{40}^{70} F(t|l = 1, m = 1, r = 2, s = 2, w)\, g(t|40 \le t < 70, w)\, dt \\
&\quad = \sum_{t=40}^{69} \int_t^{t+1} F(s|l = 1, m = 1, r = 2, s = 2, w)\, g(s|40 \le s < 70, w)\, ds \\
&\quad \ge \sum_{t=40}^{69} F(t|l = 1, m = 1, r = 2, s = 2, w) \int_t^{t+1} g(s|40 \le s < 70, w)\, ds \qquad (11) \\
&\quad = \sum_{t=40}^{69} F(t|l = 1, m = 1, r = 2, s = 2, w)[G(t + 1|40 \le t < 70, w) - G(t|40 \le t < 70, w)] \\
&\quad = K
\end{aligned}$$

where $F(t|l, m, r, s, w)$ is determined by (10) and $G(t + 1|w) - G(t|w)$ is the probability that an individual of sex $w$ in the US population aged 40 to 69 is age $t$ based on 2023 US census population single-year age distribution estimates by sex.[27] The delta method[21] as implemented in SAS/STAT PROC NLMIXED was applied to the parameter estimates for the model in (9) and their estimated covariance matrix to obtain sex-specific $K$ estimates and standard errors from (11).

**eTable 1.** Model fit for the probability of PPCM in the population of female probands with specified characteristics at a particular US advanced heart failure program

| Parameter[a] | Estimate | Standard Error | Odds Ratio (95% CI) | P-value |
|---|---|---|---|---|
| Fixed Effects | | | | |
| Intercept | -4.2149 | 0.6960 | - | - |
| Genomic ancestry | | | | |
| African | 0.2164 | 0.4235 | 1.24 (0.54 – 2.85) | 0.61 |
| European | 0 | - | - | - |
| Ethnicity | | | | |
| Hispanic | 0.8223 | 0.6516 | 2.28 (0.64 – 8.16) | 0.21 |
| Non-Hispanic | 0 | - | - | - |
| Enrollment age | | | | |
| I: [20.01, 43.15] | 3.3800 | 0.7483 | 29.37 (6.78 – 127.32) | <0.001 |
| II: [43.27, 54.11] | 2.2765 | 0.7862 | 9.74 (2.09 – 45.49) | 0.004 |
| III: [54.12, 62.78] | 1.4244 | 0.9875 | 4.16 (0.60 – 28.79) | 0.15 |
| IV: [62.92, 85.12] | 0 | - | - | - |
| Variance Component | | | | |
| Site ($\sigma^2_{\text{site}}$) | 0.07817 | 0.1432 | - | - |

CI = confidence interval.

[a] Estimated parameters for the logistic mixed model in (1) were obtained using residual subject-specific pseudolikelihood. Bias-corrected robust standard errors were obtained using the Morel-Bokossa-Neerchal correction with sites as independent units. The odds ratios and their 95% CIs, which were calculated by exponentiating corresponding estimates on the model scale, represent the change in odds of PPCM between two probands at the same site. Two-sided p-values and Wald 95% confidence intervals were calculated using the standard normal distribution.

**eTable 2.** Variants classified as P, LP, or VUS, by gene and PPCM classification of female proband(s) in which they were observed

| Gene | PPCM[a] N = 49 | | DCM[a] N = 311 | | Variants Observed in Both Classifications[a] N = 5 | | Total N = 365 | |
|---|---|---|---|---|---|---|---|---|
| N<br>% column | P/LP N = 10 | VUS N = 39 | P/LP N = 64 | VUS N = 247 | P/LP N = 2 | VUS N = 3 | P/LP N = 76 | VUS N = 289 |
| *ABCC9* | 0 | 1 | 0 | 2 | 0 | 0 | 0 | 3 |
| | 0.0 | 2.6 | 0.0 | 0.8 | 0.0 | 0.0 | 0.0 | 1.0 |
| *ACTC1* | 0 | 1 | 0 | 0 | 0 | 0 | 0 | 1 |
| | 0.0 | 2.6 | 0.0 | 0.0 | 0.0 | 0.0 | 0.0 | 0.4 |
| *ACTN2* | 0 | 0 | 0 | 2 | 0 | 0 | 0 | 2 |
| | 0.0 | 0.0 | 0.0 | 0.8 | 0.0 | 0.0 | 0.0 | 0.7 |
| *ANKRD1* | 0 | 0 | 0 | 3 | 0 | 0 | 0 | 3 |
| | 0.0 | 0.0 | 0.0 | 1.2 | 0.0 | 0.0 | 0.0 | 1.0 |
| *BAG3* | 0 | 0 | 2 | 3 | 0 | 0 | 2 | 3 |
| | 0.0 | 0.0 | 3.1 | 1.2 | 0.0 | 0.0 | 2.6 | 1.0 |
| *CRYAB* | 0 | 0 | 0 | 0 | 0 | 0 | 0 | 0 |
| | 0.0 | 0.0 | 0.0 | 0.0 | 0.0 | 0.0 | 0.0 | 0.0 |
| *CSRP3* | 0 | 0 | 0 | 2 | 0 | 0 | 0 | 2 |
| | 0.0 | 0.0 | 0.0 | 0.8 | 0.0 | 0.0 | 0.0 | 0.7 |
| *DES* | 0 | 1 | 0 | 2 | 0 | 0 | 0 | 3 |
| | 0.0 | 2.6 | 0.0 | 0.8 | 0.0 | 0.0 | 0.0 | 1.0 |
| *DSG2* | 0 | 3 | 0 | 5 | 0 | 0 | 0 | 8 |

| | | | | | | | | |
|---|---|---|---|---|---|---|---|---|
| | 0.0 | 7.7 | 0.0 | 2.0 | 0.0 | 0.0 | 0.0 | 2.8 |
| *DSP* | 0 | 6 | 5 | 22 | 0 | 1 | 5 | 29 |
| | 0.0 | 15.4 | 7.8 | 8.9 | 0.0 | 33.3 | 6.6 | 10.0 |
| *EYA4* | 0 | 1 | 0 | 2 | 0 | 0 | 0 | 3 |
| | 0.0 | 2.6 | 0.0 | 0.8 | 0.0 | 0.0 | 0.0 | 1.0 |
| *FLNC* | 1 | 2 | 13 | 22 | 0 | 1 | 14 | 25 |
| | 10.0 | 5.1 | 20.3 | 8.9 | 0.0 | 33.3 | 18.4 | 8.7 |
| *ILK* | 0 | 0 | 0 | 0 | 0 | 0 | 0 | 0 |
| | 0.0 | 0.0 | 0.0 | 0.0 | 0.0 | 0.0 | 0.0 | 0.0 |
| *JPH2* | 0 | 1 | 0 | 4 | 0 | 0 | 0 | 5 |
| | 0.0 | 2.6 | 0.0 | 1.6 | 0.0 | 0.0 | 0.0 | 1.7 |
| *LAMA4* | 0 | 2 | 0 | 13 | 0 | 0 | 0 | 15 |
| | 0.0 | 5.1 | 0.0 | 5.3 | 0.0 | 0.0 | 0.0 | 5.2 |
| *LDB3* | 0 | 3 | 0 | 12 | 0 | 0 | 0 | 15 |
| | 0.0 | 7.7 | 0.0 | 4.9 | 0.0 | 0.0 | 0.0 | 5.2 |
| *LMNA* | 1 | 0 | 6 | 5 | 0 | 0 | 7 | 5 |
| | 10.0 | 0.0 | 9.4 | 2.0 | 0.0 | 0.0 | 9.2 | 1.7 |
| *MYBPC3* | 0 | 1 | 0 | 20 | 0 | 1 | 0 | 22 |
| | 0.0 | 2.6 | 0.0 | 8.1 | 0.0 | 33.3 | 0.0 | 7.6 |
| *MYH6* | 0 | 4 | 0 | 19 | 0 | 0 | 0 | 23 |
| | 0.0 | 10.3 | 0.0 | 7.7 | 0.0 | 0.0 | 0.0 | 8.0 |
| *MYH7* | 0 | 1 | 1 | 18 | 0 | 0 | 1 | 19 |
| | 0.0 | 2.6 | 1.6 | 7.3 | 0.0 | 0.0 | 1.3 | 6.6 |

| | | | | | | | | |
|---|---|---|---|---|---|---|---|---|
| *MYPN* | 0 | 2 | 0 | 4 | 0 | 0 | 0 | 6 |
| | 0.0 | 5.1 | 0.0 | 1.6 | 0.0 | 0.0 | 0.0 | 2.1 |
| *NEBL* | 0 | 1 | 0 | 8 | 0 | 0 | 0 | 9 |
| | 0.0 | 2.6 | 0.0 | 3.2 | 0.0 | 0.0 | 0.0 | 3.1 |
| *NEXN* | 0 | 0 | 0 | 6 | 0 | 0 | 0 | 6 |
| | 0.0 | 0.0 | 0.0 | 2.4 | 0.0 | 0.0 | 0.0 | 2.1 |
| *PDLIM3* | 0 | 0 | 0 | 3 | 0 | 0 | 0 | 3 |
| | 0.0 | 0.0 | 0.0 | 1.2 | 0.0 | 0.0 | 0.0 | 1.0 |
| *PKP2* | 0 | 0 | 0 | 11 | 0 | 0 | 0 | 11 |
| | 0.0 | 0.0 | 0.0 | 4.5 | 0.0 | 0.0 | 0.0 | 3.8 |
| *PLN* | 0 | 0 | 0 | 0 | 0 | 0 | 0 | 0 |
| | 0.0 | 0.0 | 0.0 | 0.0 | 0.0 | 0.0 | 0.0 | 0.0 |
| *RBM20* | 1 | 3 | 1 | 11 | 0 | 0 | 2 | 14 |
| | 10.0 | 7.7 | 1.6 | 4.5 | 0.0 | 0.0 | 2.6 | 4.8 |
| *SCN5A* | 0 | 1 | 0 | 17 | 0 | 0 | 0 | 18 |
| | 0.0 | 2.6 | 0.0 | 6.9 | 0.0 | 0.0 | 0.0 | 6.2 |
| *SGCD* | 0 | 0 | 0 | 1 | 0 | 0 | 0 | 1 |
| | 0.0 | 0.0 | 0.0 | 0.4 | 0.0 | 0.0 | 0.0 | 0.4 |
| *TCAP* | 0 | 0 | 1 | 2 | 0 | 0 | 1 | 2 |
| | 0.0 | 0.0 | 1.6 | 0.8 | 0.0 | 0.0 | 1.3 | 0.7 |
| *TNNC1* | 0 | 1 | 0 | 2 | 0 | 0 | 0 | 3 |
| | 0.0 | 2.6 | 0.0 | 0.8 | 0.0 | 0.0 | 0.0 | 1.0 |
| *TNNI3* | 0 | 1 | 0 | 2 | 0 | 0 | 0 | 3 |

| | | | | | | | | |
|---|---|---|---|---|---|---|---|---|
| | 0.0 | 2.6 | 0.0 | 0.8 | 0.0 | 0.0 | 0.0 | 1.0 |
| *TNNT2* | 0 | 1 | 2 | 6 | 0 | 0 | 2 | 7 |
| | 0.0 | 2.6 | 3.1 | 2.4 | 0.0 | 0.0 | 2.6 | 2.4 |
| *TPM1* | 0 | 0 | 0 | 1 | 0 | 0 | 0 | 1 |
| | 0.0 | 0.0 | 0.0 | 0.4 | 0.0 | 0.0 | 0.0 | 0.4 |
| *TTN* | 7 | 2 | 33 | 14 | 2 | 0 | 42 | 16 |
| | 70.0 | 5.1 | 51.6 | 5.7 | 100.0 | 0.0 | 55.3 | 5.5 |
| *VCL* | 0 | 0 | 0 | 3 | 0 | 0 | 0 | 3 |
| | 0.0 | 0.0 | 0.0 | 1.2 | 0.0 | 0.0 | 0.0 | 1.0 |

DCM = dilated cardiomyopathy; LP = likely pathogenic; P = pathogenic; PPCM = peripartum cardiomyopathy; VUS = variant of uncertain significance.
[a] PPCM or DCM variants were observed exclusively in probands with the corresponding PPCM classification; variants identified in probands from both classifications are shown in the Variants Observed in Both Classifications column.

**eTable 3.** Characteristics of variants classified as P, LP, or VUS observed in female probands by PPCM classification

| **Variant Characteristics** | **PPCM[a] Variants N = 49** | **DCM[a] Variants N = 311** | **Variants Observed in Both Classifications N = 5** | **P-value[b]** |
|---|---|---|---|---|
| Classified P or LP, No. (%) | 10 (20.4) | 64 (20.6) | 2 (40.0) | 0.70 |
| ClinGen gene classification | | | | 0.81 |
| *TTN* (definitive) | 9 (18.4) | 47 (15.1) | 2 (40.0) | |
| Non-*TTN* definitive or strong evidence[c] | 19 (38.8) | 138 (44.4) | 2 (40.0) | |
| Moderate evidence[d] | 3 (6.1) | 18 (5.8) | 0 (0.0) | |
| Other[e] | 18 (36.7) | 108 (34.7) | 1 (20.0) | |
| Predicted impact | | | | 0.86 |
| Loss-of-function | 10 (20.4) | 74 (23.8) | 2 (40.0) | |
| Missense | 37 (75.5) | 221 (71.1) | 3 (60.0) | |
| Other | 2 (4.1) | 16 (5.1) | 0 (0.0) | |
| Band of *TTN* variants, No. (%) | [N = 9] | [N = 47] | [N = 2] | 0.81 |
| A | 8 (88.9) | 32 (68.1) | 2 (100.0) | |
| I | 1 (11.1) | 11 (23.4) | 0 (0.0) | |
| M | 0 (0.0) | 2 (4.3) | 0 (0.0) | |
| Z | 0 (0.0) | 2 (4.3) | 0 (0.0) | |

| | | | | |
|---|---|---|---|---|
| Maximum alternate allele frequency in gnomAD non-founder populations, %, median (IQR) | 0.025 (0.008 - 0.059) [N = 31] | 0.012 (0.004 - 0.044) [N = 221] | 0.055 (0.021 - 0.104) [N = 4] | 0.02 |
| Absent in non-founder gnomAD populations, No. (%) | 18 (36.7) | 90 (28.9) | 1 (20.0) | 0.54 |
| REVEL score for missense variants, median (IQR) | 0.50 (0.23 - 0.77) [N = 37] | 0.47 (0.25 - 0.71) [N = 220] | 0.36 (0.24 - 0.61) [N = 3] | 0.79 |
| Classification criterion met, No. (%)[f] | | | | |
| Population frequency domain | | | | 0.35 |
| Absent from gnomAD or rare (<0.05%) in all gnomAD nonfounder populations with available data (PM2) | 32 (65.3) | 226 (72.7) | 2 (40.0) | |
| Variant allele frequency is >0.05% in any gnomAD nonfounder population (BS1) | 3 (6.1) | 19 (6.1) | 1 (20.0) | |
| None of PM2, BS1, or BA1 | 14 (28.6) | 66 (21.2) | 2 (40.0) | |
| Null variant in gene/region | | | | 0.53 |
| Null variant in *LMNA* or *SCN5A* (PVS1) | 1 (2.0) | 3 (1.0) | 0 (0.0) | |
| Null variant in *FLNC*, *BAG3*, *DSP*, or *TTN* (PVS1_Strong) | 8 (16.3) | 53 (17.0) | 2 (40.0) | |
| Null variant in *VCL* or, *PLN*, (PVS1_Moderate) | 0 (0.0) | 0 (0.0) | 0 (0.0) | |
| None of PVS1, PVS1_Strong, or PVS1_Moderate | 40 (81.6) | 255 (82.0) | 3 (60.0) | |
| Segregation domain | | | | 0.02 |

| | | | | |
|---|---|---|---|---|
| Variant segregates with disease in ≥7 meioses in DCM genes with established evidence (PP1_Strong) | 0 (0.0) | 5 (1.6) | 0 (0.0) | |
| Variant segregates with disease in ≥5 meioses in DCM genes with established evidence (PP1_Moderate) | 0 (0.0) | 1 (0.3) | 1 (20.0) | |
| Variant segregates with disease in ≥3 meioses in DCM genes with established evidence (PP1) | 0 (0.0) | 3 (1.0) | 0 (0.0) | |
| None of PP1_Strong, PP1_Moderate, PP1, or BS4 | 49 (100.0) | 302 (97.1) | 4 (80.0) | |
| Case counts or reported association domain | | | | 0.08 |
| Prevalence of variant in affected individuals is significantly increased compared with controls OR variant identified in ≥15 probands with consistent phenotypes OR variant identified in ≥10 confirmed unrelated probands with consistent phenotypes (PS4) | 0 (0.0) | 5 (1.6) | 1 (20.0) | |
| Variant identified in ≥6 probands with consistent phenotypes (PS4_Moderate) | 0 (0.0) | 8 (2.6) | 0 (0.0) | |
| Variant identified in ≥2 probands with consistent phenotypes (PS4_Supporting) | 7 (14.3) | 57 (18.3) | 1 (20.0) | |
| None of PS4, PS4_Moderate, or PS4_Supporting | 42 (85.7) | 241 (77.5) | 3 (60.0) | |
| *In silico* predictions domain | | | | 0.34 |
| Multiple lines of computational evidence support a deleterious effect (REVEL score >0.7; PP3) | 14 (28.6) | 62 (19.9) | 0 (0.0) | |

| | | | | |
|---|---|---|---|---|
| Multiple lines of computational evidence suggest no impact on gene or gene product (REVEL score <0.15; BP4) | 3 (6.1) | 28 (9.0) | 0 (0.0) | |
| Neither PP3 nor BP4 | 32 (65.3) | 221 (71.1) | 5 (100.0) | |
| Functional studies domain | | | | 1.00 |
| Well-established in vitro or in vivo functional studies supportive of a damaging effect (PS3) | 0 (0.0) | 2 (0.6) | 0 (0.0) | |
| Well established in vitro or in vivo functional studies show no damaging effect (BS3) | 0 (0.0) | 0 (0.0) | 0 (0.0) | |
| Neither PS3 nor BS3 | 49 (100.0) | 309 (99.4) | 5 (100.0) | |
| Missense change domain | | | | 0.38 |
| Different missense change at same amino acid residue previously established as pathogenic (PM5) | 1 (2.0) | 2 (0.6) | 0 (0.0) | |
| Neither PS1 nor PM5 | 48 (98.0) | 309 (99.4) | 5 (100.0) | |
| De novo domain | | | | 0.27 |
| De novo (paternity confirmed) in a patient with disease and no family history (PS2) | 0 (0.0) | 0 (0.0) | 0 (0.0) | |
| De novo without confirmation of paternity (parental clinical data required; PM6) | 1 (2.0) | 1 (0.3) | 0 (0.0) | |
| Neither PS2 nor PM6 | 48 (98.0) | 310 (99.7) | 5 (100.0) | |

| | | | | |
|---|---|---|---|---|
| Located in hotspot (for DCM, RBM20 exon 9, amino acids 634, 636, 637, 638; PM1) | 1 (2.0) | 1 (0.3) | 0 (0.0) | 0.27 |
| In-frame deletions/insertions of any size in a nonrepeat region or stop-loss variants (PM4) | 2 (4.1) | 17 (5.5) | 0 (0.0) | 0.82 |
| Observed as a compound heterozygote (in trans) or double heterozygote in genes with overlapping function (BP2) | 0 (0.0) | 0 (0.0) | 0 (0.0) | - |

DCM = dilated cardiomyopathy; IQR = interquartile range; LP = likely pathogenic; P = pathogenic; PPCM = peripartum cardiomyopathy; VUS = variant of uncertain significance.

[a] PPCM and DCM variants were observed exclusively in probands with the corresponding classification; variants identified in probands from both classifications are shown in the Variants Observed in Both Classifications column.

[b] Variant characteristics were compared across ancestries using the exact Pearson chi-square test for nominal variables and the Kruskal-Wallis test for continuous variables. The exact Monte Carlo p-value estimate based on 100,000 replicates was reported for the Pearson chi-square test. Post-hoc pairwise comparisons between ancestries for $p<0.05$ are presented in eTable 4.

[c] Non-*TTN* definitive or strong genes by ClinGen classification that were included in this study were *BAG3*, *DES*, *DSP*, *FLNC*, *LMNA*, *MYH7*, *PLN*, *RBM20*, *SCN5A*, *TNNC1*, and *TNNT2*.

[d] Non-*TTN* moderate genes by ClinGen classification that were included in this study were *ACTC1*, *ACTN2*, *JPH2*, *NEXN*, *TNNI3*, *TPM1*, and *VCL*.

[e] Other genes included *ABCC9*, *ANKRD1*, *CRYAB*, *CSRP3*, *DSG2*, *EYA4*, *ILK*, *LAMA4*, *LDB3*, *MYBPC3*, *MYH6*, *MYPN*, *NEBL*, *PDLIM3*, *PKP2*, *SGCD*, and *TCAP*.

[f] Mutually exclusive criteria relating to a single domain (e.g., population frequency) were analyzed as a multinomial outcome for that domain. Criteria not met for any variant that was manually classified, which included BA1 (population frequency domain), BPA1, BP7, PS1 (missense change domain), PM3, and BS4 (segregation domain), are not shown in the table.

**eTable 4.** Post-hoc pairwise comparisons of variant characteristics that differed across PPCM classifications with P<0.05

| Variant Characteristics | Variants Observed in Probands of… | Compared to… | P-value[a] |
|---|---|---|---|
| Maximum alternate allele frequency in gnomAD non-founder populations | PPCM | DCM | 0.04 |
| | PPCM | Both PPCM and DCM | 0.68 |
| | DCM | Both PPCM and DCM | 0.32 |
| Segregation domain | PPCM | DCM | 0.68 |
| | PPCM | Both PPCM and DCM | 0.19 |
| | DCM | Both PPCM and DCM | 0.11 |

DCM = dilated cardiomyopathy; PPCM = peripartum cardiomyopathy.
[a] Holm-Bonferroni corrected exact Monte-Carlo p-values based on 100,000 replicates for the chi-square test involving only the two disease groups of interest are presented for nominal variant characteristics, while the Dwass, Steel, Critchlow-Fligner method was applied to continuous variant characteristics.

**eTable 5.** Model fit for the age-specific cumulative risk of DCM only in a first-degree relative of a female proband with PPCM or DCM

| Parameter[a] | Estimate | Standard Error | Hazard Ratio (95% CI) | P |
|---|---|---|---|---|
| Fixed Effects | | | | |
| Weibull baseline survivor function parameters[b] | | | | |
| *a* | -7.9183 | 1.6234 | - | - |
| *b* | 1.5291 | 0.4313 | - | - |
| Proband PPCM classification | | | | |
| PPCM | -0.5421 | 0.4721 | 0.58 (0.23 - 1.47) | 0.25 |
| DCM | 0 | - | 1.00 | - |
| First-degree relative sex | | | | |
| Female | -0.2696 | 0.3403 | 0.76 (0.39 - 1.49) | 0.43 |
| Male | 0 | - | 1.00 | - |
| Proband ancestry group | | | | |
| African Ancestry | 0.7794 | 0.2310 | 2.18 (1.39 - 3.43) | 0.001 |
| European Ancestry | 0 | - | 1.00 | - |
| Proband self-reported ethnicity | | | | |
| Hispanic | 0.2138 | 0.8197 | 1.24 (0.25 – 6.17) | 0.79 |
| Non-Hispanic | 0 | - | 1.00 | - |
| Proband diagnosis age | | | | |
| I: [9.78, 34.99] | 0 | - | 1.00 | - |
| II: [35.01, 44.47] | -0.01464 | 0.4027 | 0.99 (0.45 – 2.17) | 0.97 |
| III: [44.48, 54.09] | -0.3490 | 0.4795 | 0.71 (0.28 – 1.81) | 0.47 |
| IV: [54.21, 78.31] | -0.6165 | 0.5356 | 0.54 (0.19 – 1.54) | 0.25 |
| Variance Components | | | | |

| | | | | |
|---|---|---|---|---|
| Residual | 0.9532 | 0.05263 | - | - |

CI = confidence interval; DCM = dilated cardiomyopathy; PPCM = peripartum cardiomyopathy.

[a] Parameters of a Weibull proportional hazards model for age-specific cumulative risk were estimated using a generalized estimating equation-type generalized linear mixed model with a binary outcome (presence or absence of DCM), complementary log-log link, random proband enrollment site intercept, and working independence correlation structure fit using residual subject specific pseudolikelihood. Bias-corrected robust standard errors were obtained using the Morel-Bokossa-Neerchal correction with sites as independent units. The site random effect was estimated to be zero and was therefore excluded from the model. Two-sided p-values and Wald 95% confidence intervals were calculated using the standard normal distribution. Hazard ratios and their 95% confidence intervals were obtained by exponentiating corresponding estimates on the model scale.

[b] *a* and *b* are parameters of the Weibull baseline survivor function of the form $S_o(t) = \exp[-\exp(a)t^b]$. 665 first-degree relatives (103 of PPCM probands and 562 of DCM probands) contributed to this analysis.
